# COVID-19 Spreading Dynamics with Vaccination – Allocation Strategy, Return to Normalcy and Vaccine Hesitancy

**DOI:** 10.1101/2020.12.10.20247049

**Authors:** B Shayak, Mohit M Sharma

## Abstract

In this work we use mathematical modeling to analyse the dynamics of COVID-19 spread after a vaccination program is initiated. The model used is a delay differential equation developed earlier by our group. Basis of currently available data, our principal findings are as follows. (*a*) For fastest deceleration of the pandemic, people with high interaction rate such as grocers and airline cabin crew should be given priority in vaccine access. (*b*) Individuals who have been vaccinated may be selectively cleared to return to normal activities without significant risk of a resurgence in cases. (*c*) If an infection as well as a vaccine confers immunity for a duration *τ*_0_, then the pandemic can be eliminated by vaccinating people at a sufficiently high rate. Unless *τ*_0_ is very small, the cutoff rate required appears feasible to achieve in practice. (*d*) The presence of a substantial minority of vaccine-hesitant population might not amount to a significant threat or even an inconvenience to a vaccine-compliant majority population.

## §0 INTRODUCTION

Last Wednesday, on 02 December 2020, a COVID-19 vaccine candidate developed by the American company Pfizer was authorized for emergency use in the United Kingdom. Yesterday, dubbed V-Day in the British media, it was distributed to the first batch of UK citizens, thus marking the start of the endgame phase in our fight against the virus. As we write this, Pfizer’s candidate, which is based on the messenger RNA platform, is pending approval in USA, India and in the European Union. As a first vaccine, Pfizer does have some limitations, chief among them being the requirement for storage at cryogenic temperatures. Other front-running vaccines do not have this restriction however. The candidate developed by Moderna, USA, which is currently pending approval in USA, UK and Europe, has a more tractable storage requirement though it is also based on messenger RNA like the Pfizer vaccine. A different approach – that of viral vectoring or grafting the COVID-19 antigen onto a harmless adenovirus – has been employed for the vaccine candidate developed by Oxford University, UK. This vaccine is commercially manufactured by Astra Zeneca, UK for the western market and by Serum Institute of India for the eastern market, where it is known as Covishield. It is right behind Pfizer and Moderna in the queue for approval, and, requiring only conventional refrigeration for the entire time from manufacture to administration, is an especially promising candidate for distribution on a massive scale. Serum Institute has already manufactured multiple crores of doses of Covishield, and filed for approval in India on Sunday evening.

Behind the three leading candidates there are runners-up which also hold considerable promise. The ICMR-Bharat Biotech candidate called Covaxin, which is currently well into Phase 3 trials, is based on inactivated virus and represents a tried and tested technology as opposed to a new approach. Despite the last-stage trial still in progress, Bharat Biotech has filed for emergency use authorization on Monday, one day following Covishield. The Johnson and Johnson candidate, based on viral vector like Covishield, is also in Phase 3 trials and has the potential to require just a single dose, unlike the mandatory two doses of all the other candidates mentioned so far. The Russian vaccine called Sputnik 5 is also undergoing combined Phase 2/3 trials in India, though it has already been approved for use in Russia. The absence of transparency in the Russian data, combined with the fact that coronavirus cases in Russia have ballooned after introduction of the vaccine have necessitated scepticism and re-trials in the rest of the world. Sputnik may still prove effective in trials though, and without a thorough examination it cannot be discarded as a candidate. With trillions of doses being required to immunize the entire world’s population, the more manufacturers there are, the better it will be.

With a new vaccine, a critical question is its degree of efficacy. Pfizer and Moderna have both reported 90-plus percentage of efficacy for their vaccine candidates. For Covishield, the scenario is a little more complicated; in the western trials, Astra Zeneca has declared 62 percent efficacy in a group which received two “full-strength” doses of vaccine and 90 percent efficacy in a group which accidentally received a “half-strength” first dose. Serum Institute has not yet reported an efficacy factor for the eastern branch of the trials. The definition of efficacy is as follows : in Phase 3 trials, half the participants are given the vaccine while the other half are given a placebo. In the weeks following administration, all participants are monitored and COVID-19 cases cropping up among them are counted. If *n*_1_ cases occur among the placebo group and *n*_2_ among the vaccine group, then the efficacy is defined as *E* = 100 x (*n*_1_ − *n*_2_)/*n*_1_ percent. A fully effective vaccine means zero cases in the vaccine group or *n*_2_ = 0, which leads to *E* = 100. A completely ineffective vaccine means equal numbers of cases in the vaccine and the placebo group or *n*_1_ = *n*_2_ which leads to *E* = 0. This is a more indirect definition than that obtained from a hypothetical challenge experiment where vaccinated people are deliberately exposed to the virus and the number of infections is counted. Such an experiment is obviously unethical however so we must make do with the efficacy as defined above.

An equally important index for a vaccine is the number of COVID-19 hospitalizations and deaths arising among the vaccine group. That is, even if the vaccine cannot outright prevent contraction of the disease, can it at least ensure that the disease does not progress to the severe state. In an epidemic like COVID-19 where mild disease states are norm rather than exception, a vaccine which causes more cases but ensures this state for all recipients is more useful than one which causes fewer cases but these include hospitalizations and/or deaths. So far, Pfizer has reported one incidence of hospitalization among the vaccine group while Moderna and Covishield have reported zero. Of course, the numbers currently involved are way too small to make any kind of quantitative deduction regarding the effectivity of the different vaccines at preventing severe disease. A second possible partial action of a vaccine is in reducing transmissibility – a vaccinated corona patient might spread the disease to less people than an unvaccinated one. In our view, this is a less important endpoint than severity reduction, especially from the general public’s viewpoint – if I take a vaccine, I will be more concerned about whether I will contract the disease and whether I will end up in hospital than about who else I transmit the virus to.

With the release of the first vaccine, the conquest of the pathogen by modern science and technology is clearly in sight. That happy event hasn’t arrived yet though. Just today, two lakhs of new coronavirus cases have cropped up in USA, multiple thousands each in western European countries, 40,000 in Brazil, 30,000 in India and so on. Even with all vaccine factories running full throttle, the doses will flow out to the world’s population like a leaky faucet dripping into a bucket. Some questions immediately arise. How long will it be before the Armistice arrives ? How should we apportion the limited doses so that it arrives the quickest ? When it does come, will it be an unconditional surrender or an uneasy truce ? How hard can the virus harass us before giving up the ghost ? There is only one approach to answering questions like this, which is mathematical modeling. Let’s get down to work without further ado.

## §1 INTERACTION-STRUCTURED DDE MODEL

The model we use for the analysis is a delay differential equation (DDE) developed by our group [1]. On account of the novelty of the model, we present here a very quick summary. The model operates in a region such as a town, village or neighbourhood with good interaction among its inhabitants. It uses a single dependent variable, *y* (*t*), which is the cumulative count of corona cases in the region as a function of time. It treats disease transmission as the result of interaction, either inter-personal or via objects (fomites), between at large cases and targets (susceptible people to whom the case transfers the virus). Its philosophy is described by the word equation

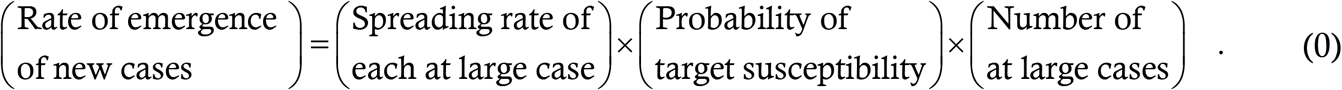

The left hand side (LHS) of this equation is d*y*/d*t*. The first term on the right hand side (RHS) is the product of two things : the rate (persons/day) at which an at large case interacts with other people and the probability that an interaction with a susceptible target actually leads to transmission. The second term on the RHS accounts for the immune response. The third term counts the number of at large cases. It depends on the following parameters :

- *μ*_1_ : the fraction of cases who are asymptomatic
- *μ*_3_ : the fraction of cases (both asymptomatic and symptomatic) who do NOT get caught in contact tracing drives
- *τ*_1_ : the asymptomatic infection period i.e. the duration for which untraced asymptomatic patients remain transmissible and at large
- *τ*_2_ : the latency period i.e. the duration for which untraced symptomatic patients remain transmissible and at large before manifesting symptoms and going into quarantine
- *T* : the duration for which contact traced cases (both asymptomatic and symptomatic) remain transmissible and at large before being caught and quarantined

For any lumped parameter or compartmental model, the parameter values used must be averaged over all the cases. It can be shown [1] that the mathematical representation of the third term on the RHS of (0) is

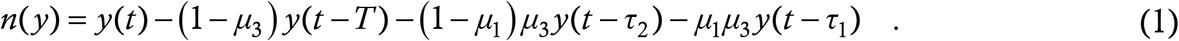

This acts as the definition of the function *n*(·), a useful shorthand for a cumbersome but conteptually simple expression; it consists of delay terms, which is why the model is a DDE.

In Ref. [1], we have taken the baseline version of the model to be one where a bout of infection confers lifetime immunity. This leads to the second term on the RHS of (0) acquiring the form 1 − *y*/*N*, where *N* is the total number of susceptible people present in the region at the start of the outbreak. Finally, we have defined the per-case spreading rate to be *m*_0_ to get the equation

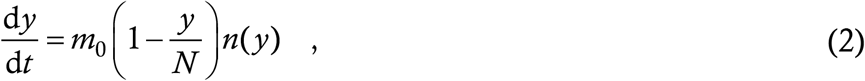

which is the **retarded logistic equation**. The reproduction number *R* at any stage of the disease evolution can be calculated from this model in a very transparent form; the expression turns out to be

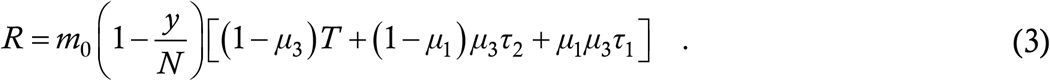

When *y* = 0, *R* equals the basic value *R*_0_. This completes the recapitulation of the prior work. As we have already mentioned, this is just a summary and for more details we must refer you to Ref. [1]. We have chosen this model instead of another one for the present analysis because (*a*) all parameters in the model are directly related to the disease and its management instead of being heuristic or ad hoc, (*b*) the model has displayed excellent accuracy in predicting the trajectories of the virus in different parts of the world and *(c)* it can be easily adapted to accommodate different kinds of situations. Again, these points have been elaborated in Ref. [1] with a variety of examples.

To model vaccination, an obvious step we have to take is the introduction of a vaccinated class of people. Before that however, we shall make an extension of the baseline model (2) to accommodate a feature of real society which is very important. This is that not everyone has the same rate of interaction with other people. In a full or partial lockdown, which the whole world is currently under, this is all the more true. An essential worker such as a grocer or a waiter has no option but to interact with customers all day. On the other hand, a person such as a working from home software engineer might have everything delivered to his/her house and have a negligible interaction rate. It is often said that 80 percent of the secondary cases are spawned by 20 percent of the primary ones, or similar statements with the numbers somewhat changed. This phenomenon arises because of the heterogeneous interaction rate. It turns out that this has significant implications for vaccination strategies, so we shall build interaction-structuring into the model from the outset.

Here we use a **two-component model**. Let there be *N*_1_ people who have very high interaction rate and *N*_2_ people with *N*_1_+*N*_2_ = *N* who have a lower interaction rate. Since the spreading rate *m*_0_ in (2) is proportional to the interaction rate, let *m*_1_ be the per-case spreading rate for the high-interaction class and *m*_2_ be that for the low-interaction class. We assume that every person interacts equally with people of both classes i.e. a fraction *N*_1_/*N* of any person’s interactions will be with high-interaction people and a fraction *N*_2_/*N* of these interactions will be with low-interaction people. Letting *y* denote the cumulative case count in the high-interaction class and *z* that in the low-interaction class, the two-component extension of (2) reads as

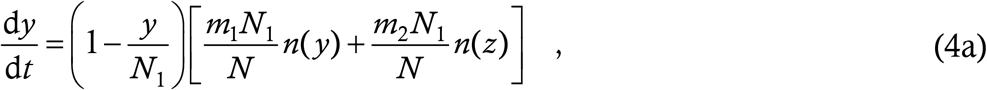

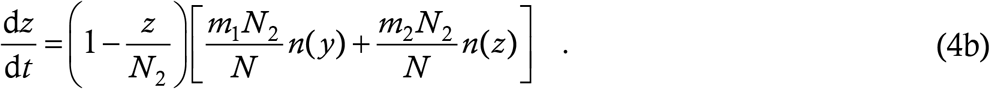

The factor *N*_1_/*N* in the RHS of (4a) denotes the probability, mentioned above, that the target of the interaction is a high-interaction person, and similarly for (4b). For more details of the derivation of structured equations within our DDE framework, please consult §5 of Ref. [1]. From (4), it should be clear how to generalize the model to include even more interaction classes. In this Article, we shall not use more than two classes.

Finally, the vaccine itself gets the cue to walk onto the stage. Let *v* denote the cumulative number of high-interaction people who have been vaccinated and *w* denote the cumulative number of low-interaction people who have been vaccinated. We make the following postulates and assumptions for the model.

- A person **switches instantly** rather than gradually from non-vaccinated to vaccinated. For the most realistic scenario with a two-dose vaccine, the instant of transition will probably be somewhere between the first and second doses. To model a worst-case scenario, one can treat this instant to be after the second dose. Technically, the vaccination point is defined as a prescribed number of days following the second dose. A high-profile corona case has occurred in Haryana, India, featuring a minister who contracted the disease two weeks after receiving the first dose of Covaxin in a Phase 3 trial (the second dose is due four weeks after the first). Technically, this case shall not be considered as an instance of ineffectivity of the vaccine since for Covaxin the vaccinee is considered immune only after 14 days following the second dose.
- If a person is vaccinated, there is a probability *η* that s/he will not contract the disease when exposed to the virus. This *η*, which we call the **effectivity**, may be different from the published efficacy *E* mentioned in §0 due to the difference between the definitions of *E* and *η*. In §4 we shall analyse the relationship between *E* and *η*. We assume that all administered vaccines have the same *η*.
- If a vaccinated person contracts the disease, then s/he will have the **same transmission properties** (*μ*_1_, *τ*_1_ etc) as an unvaccinated person who contracts it i.e. *n*(·) will feature the same parameters for both vaccinated and unvaccinated cases. It may be true that vaccinated cases are less transmissible; that can be easily taken care of if the data suggest the need to do it.
- Vaccines are manufactured at a constant rate *α* per day. A fraction *β* of these shots is allocated to the high-interaction class and the rest to the low-interaction class. People who contract COVID-19 before being vaccinated are treated as immune and not given the vaccine following recovery. If all people of one class have either caught COVID or been vaccinated, all the *α* vaccines per day are distributed to the other class. When that class has been saturated as well, the vaccination drive ceases.

The last postulate leads to the following equations for the vaccinated people *v* and *w* :

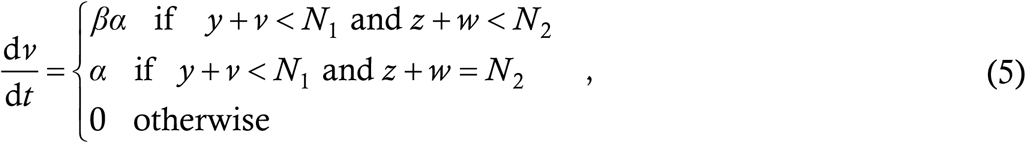

and by analogy

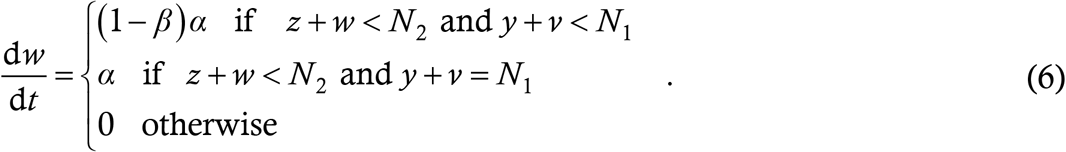

These equations implicitly assume that everyone is willing to take the vaccine i.e. there is no vaccine hesitancy. We shall relax this assumption in a later Section. With partial vaccine effectivity, Equations (5,6) also contain another simplification. The stopping condition *y* + *v* = *N*_1_ (and the equivalent for *N*_2_) assumes that everyone is either a case or a vaccinee. With limited vaccine effectivity however, the two sets can have a non-trivial overlap. In other words, a subset say *u* of the cases *y* will actually have occurred among the vaccine group *v*, and the stopping criterion (5) will then deprive *u* healthy susceptible people of the vaccines. With high effectivity however, the error involved is small. Correcting it involves expansion of the two-component model to a four-component one, a procedure we shall describe and undertake in the later Sections. Since the simplification results in some people missing out on the vaccine, the simplified model should generate **higher** case counts than a more accurate four-component model. We feel that for a basic model, this extra complexity is not worth undertaking, especially since the errors here will drive us towards more cautious predictions and policy recommendations.

Given the equations for *v* and *w* let us now hack the math on *y* and *z*. We start with *y* first. Without vaccines, the structure (0) leads to the equation (4a). By our assumptions, *n*(*y*) and *n*(*z*) are the same for cases who have received vaccine or otherwise. For the basic model, the interaction rates *m*_1_ and *m*_2_ also remain the same for vaccinated and unvaccinated people. The only thing which changes is the susceptibility probability. If a person has received the vaccine, then there is a probability *η* that s/he is insusceptible. For the basic model, we assume that infection as well as vaccine render lifetime immunity (later we will deal with the case where this is not true). To calculate the susceptibility probability in (0), we first count the insusceptibles in the high-interaction class. These are all those who have contracted the disease, and fraction *η* of those who have received the vaccine i.e. the total number of insusceptibles is *y*+*ηv*. Thus, the probability that a random high-interaction person is insusceptible is (*y*+*ηv*)/*N*_1_ and the probability of susceptibility is 1 − (*y*+*ηv*)/*N*_1_. Plugging this into the structure (0) yields

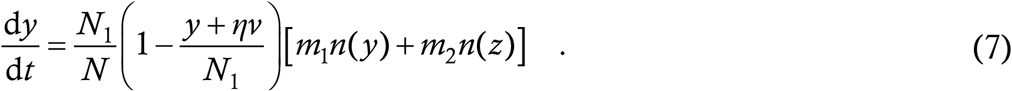

The term *N*_1_/*N* is just brought to front from the box bracket in (3a). Repeating the logic for the low-interaction class leads to

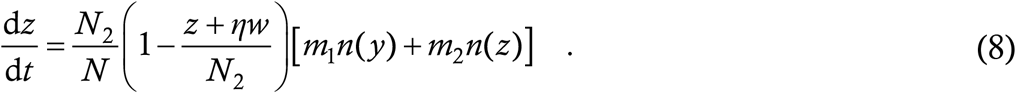

Equations (5-8) constitute the basic two-component interaction-structured coronavirus model with vaccination.

The procedure we shall use to solve this model as well as the variants considered in later Sections will be numerical integration in Matlab using 2^nd^ order Runge Kutta method with a step size of *h* = 1/1000 day. We shall keep some features **constant for all the runs** i.e. throughout this Article. Among these invariants are the parameter values *μ*_1_ = 0·8, *μ*_3_ = 1, *τ*_1_ = 7 and *τ*_2_ = 3. The assumption of no contact tracing might appear surprising. However, we have shown in §6 of Ref. [1] that, with high fraction of asymptomatic carriers, the percentage of cases who are caught is often frustratingly small. Moreover, contact tracing drives are currently managed by healthcare workers; once a vaccine arrives, many of them will be redeployed to vaccination drives and the tracing efforts will further lose steam. Finally, we have observed in runs of the baseline model that the values of the specific parameters in *n*(·) are not of the greatest importance, in that the solution curves are very similar if the parameters are changed in such a way as to keep *R*_0_ a constant. For each run in this Article, we shall solve the model in a Notional City having an initial susceptible population of *N* = 3,00,000. This is the same solution domain as has been used in the prior work [1]. For all runs we shall assume that there are zero pre-existing cases when the vaccination drive begins, and seed the model with a very small number of cases – typically constant 10 cases/day of each kind for seven days. Accounting for the initial presence of existing cases is merely algebraic trouble but adds no novel conceptual insights. The solutions are to the largest degree independent of the seeding function. At the other end, we shall declare the epidemic to be over using the following criterion – defining the active case count to be the cumulative count today less the count 14 days back, we shall terminate the epidemic if there is fewer than one active case for 14 consecutive days. This criterion has also been used by us in Ref. [2].

In what follows, we shall use the basic model (5-8) with suitable additions as needed to analyse several different scenarios which arise in the context of vaccination. We shall begin the consideration of each scenario with a question which a reader of a typical “ask the experts” column is likely to pose. We shall then use our model to attempt an answer to that question and, with the help of simulation results, highlight the various issues involved. After that, we shall include a discussion on the implications of the model results for policy makers. **The disclaimer applies throughout this Article that a mathematical model is only an approximation of reality and its predictions are NOT guaranteed to come true**.

## §2 OPTIMAL VACCINATION STRATEGY

### QUESTION : Who gets the vaccine first ?

“Not me,” is what almost everyone will be thinking. So let us rephrase the question as “When will my turn come ?” When vaccines are in limited supply, the issue of who gets it first is very important since it turns out that the priority order can have a significant impact on the time taken to stop the pandemic and the number of cases generated in the process.

Before getting into the modeling, we express our opinion, echoed by almost all governments across the world, that the first in line should be the healthcare workers, irrespective of their contribution to COVID-19 spread. This is because every medical professional including doctors, nurses, testing centre employees, hospital orderlies, cleaners, radiologists and hospital technicians have been at the frontlines of battle for the past ten months and have acted as the bulwark on which the global disease management strategy has rested. As a community, their contribution to the corona cause has far exceeded that of any other group, so they should be given the first vaccines as a reward for their efforts. Personnel at organizations like Moderna, ICMR and Oxford University, who have spent countless hours often handling live virus to develop the vaccines in record time, should also be included within the earliest priority group on account of their seminal contributions. The same holds for staff who have worked on drug development and repurposing, especially in settings involving risk. (It is also our opinion that a distant second but non-trivial contribution to the cause has come from mathematical modellers, so they should be included in the highest-but-one priority group – some people however may disagree.)

But after the medical care workers are taken care of, then who ? To answer that, we turn to the model. Here we shall use the basic model (5-8) since that already features interaction-structuring. As a reference scenario, we consider the case where 10 percent of the population has an interaction rate significantly higher than the remaining 90 percent i.e. we take *N*_1_ = 30,000 and *N*_2_ = 2,70,000. We shall refer to this choice as the **90—10 social structuring**. During a restricted mobility phase let us take *m*_1_ = 5*m*_2_; to determine their values, we turn to the reproduction number. A data fit of New York State, USA and Italy during hard lockdown [3] indicates *R*_0_ = 1·15 during such a phase. With the parameter values of §1, (3) yields *m*_0_ = 5/31 = 0·1613 for *R*_0_ to be unity.

For the first set of simulations we choose *m*_1_ = 0·75 and *m*_2_ = 0·15, which gives an *R*_0_ of approximately 1·3, slightly higher than during a hard lockdown. As a trial run, we simulate the model with no vaccination whatever, and show the results below. We plot *y* in blue and *z* in green. In place of the derivatives themselves, we show the epidemiological curve or “epi-curve” which is a bar chart of the number of cases cropping up in each week of the outbreak. So that the epi-curve may form a tangent to the derivative, we have scaled it down by a factor of 7, just as in the prior works [1,2]. The epi-curve for *y* is in grey and for *z* in cyan.

We see a nice bell-shaped epi-curve in the absence of vaccines. Although the derivative bars are no longer visible after the 160^th^ day, it actually turns out that the mathematical stopping condition as we have defined it (§1, less than one active case for 14 consecutive days) occurs at 241 days. The terminal number of cases is 1,27,600 which amounts to approximately 40 percent infection level.

We now introduce vaccination, delivering vaccines at the rate of *α* = 600 per day. This vaccinates the entire population in 500 days, which is quite reasonable. We take the vaccine effectivity as *η* = 0·8, assuming that the company estimates of efficacy are over-enthusiastic. For the first run, we take *β* = 0·1 i.e. the vaccines are allotted to the high- and low-interaction classes in proportion to their population. In addition to all the things in Fig. 1, we show below the time traces of *v* and *w* in red and magenta respectively.

**Figure 1:**
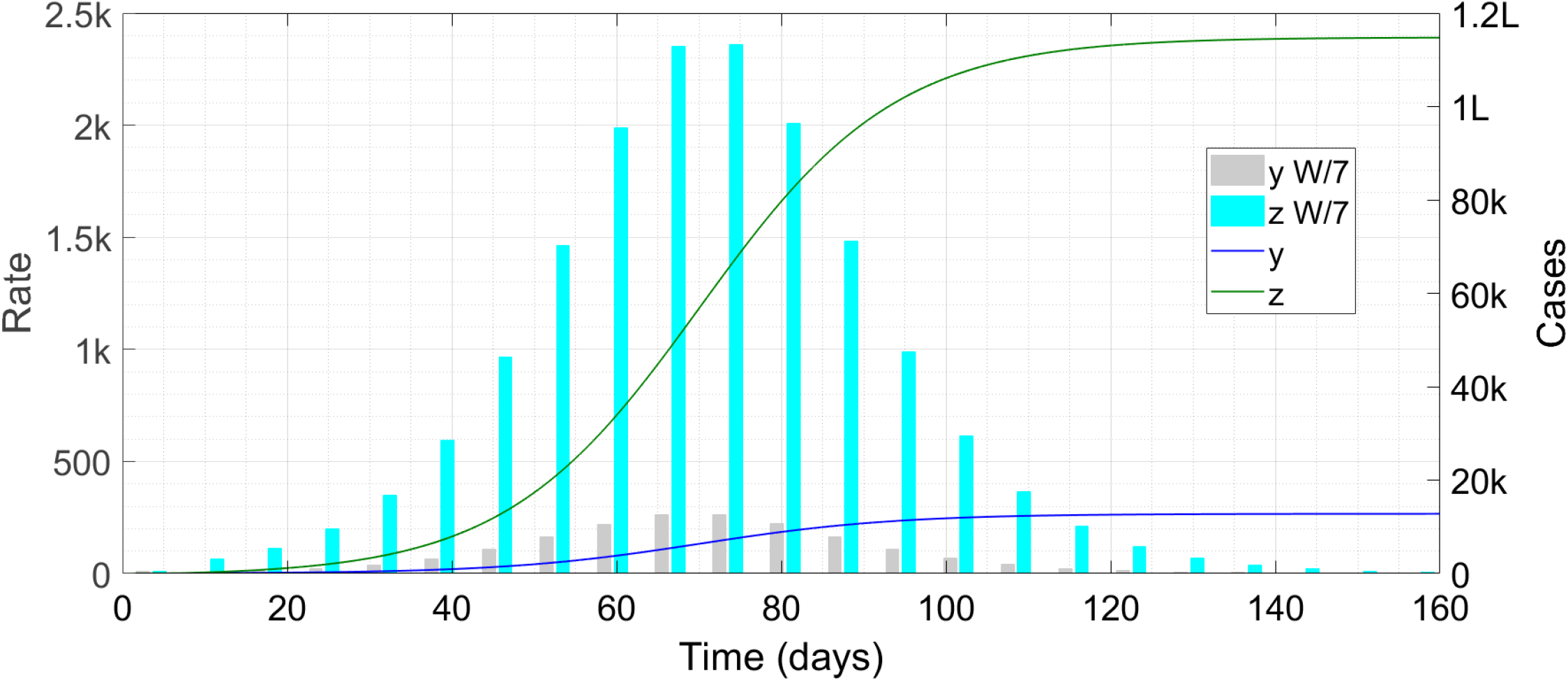
Time trace of the epidemic without vaccination. The symbol ‘k’ denotes thousand, ‘L’ hundred thousand and ‘W/7’ weekly cases scaled down by a factor of seven.

The technical time to the end of the outbreak is 214 days, by which time 71,000 cases have broken out and nearly 1,25,000 vaccines have been distributed. In this Figure, we have stopped the vaccination drive when the epidemic ceases, which is a minor detail. Note that the peak case rate has become half of what it was in Fig. 1. For the next run, we change *β* to 0·9 i.e. we now preferentially vaccinate the high-interaction class first.

This time, the very rapid initial vaccination of the high-interaction people takes the wind out of the sails of the disease. Formally, the total runtime of the disease has not shortened appreciably from the previous run with the endpoint now being 203 days. The peak however comes 20 days earlier than in Fig. 2 and the total case count has reduced by a factor of 4·5 to less than 16,000. The number of vaccines distributed upto the end is just less than 1,18,000.

**Figure 2:**
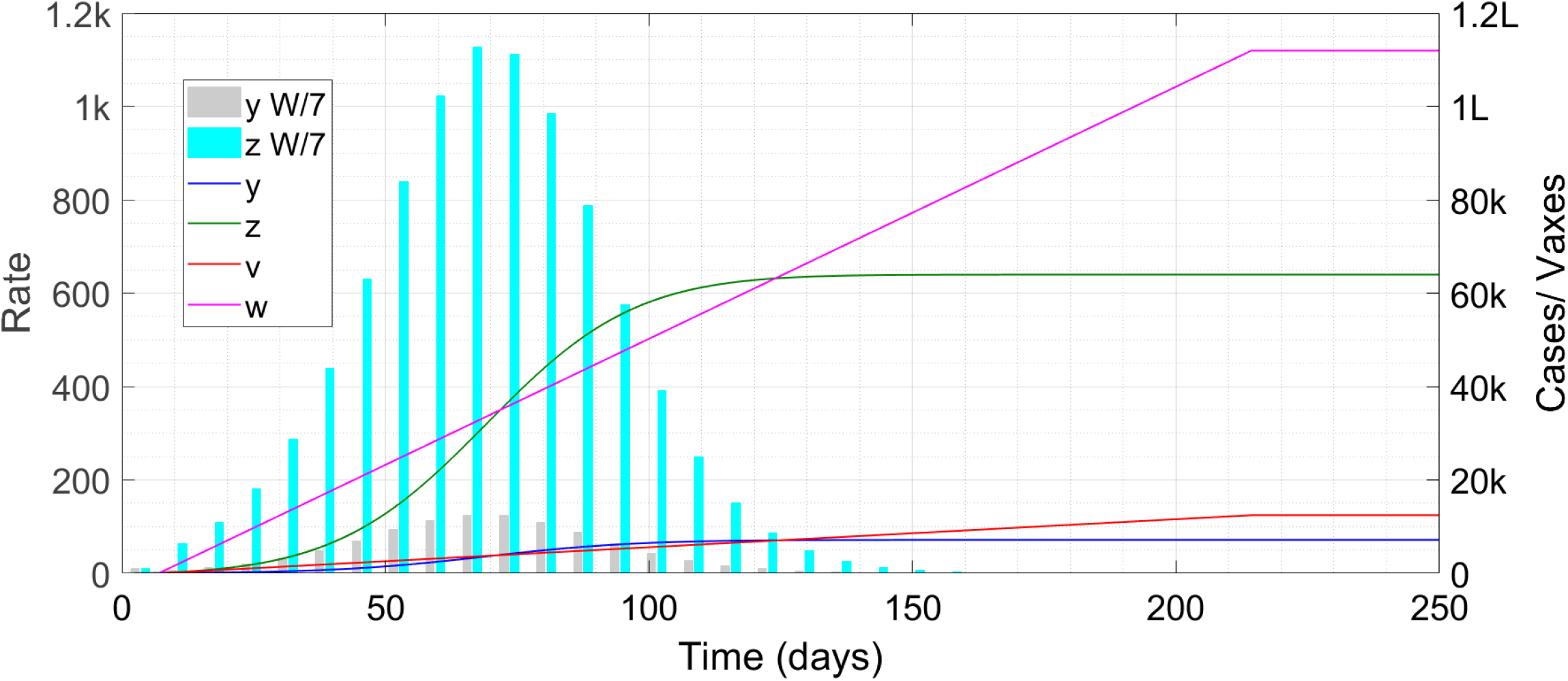
Time trace of the epidemic with vaccination. The symbol ‘k’ denotes thousand, ‘L’ hundred thousand and ‘W/7’ weekly cases scaled down by a factor of seven.

Thus, we can see that **preferential vaccination of the high-interaction class** can result in faster termination and much lower infection counts. To bolster our case, we consider a few more scenarios of this nature. We take three kinds of interaction-structure – (*a*) the present one, (*b*) a situation “95—05” where 5 percent of the population has 10 times the interaction rate of the rest and (*c*) a situation “80—20” where 20 percent of the people have 3 times the interaction rate of the rest. In the latter two situations, we use the values *m*_1_ = 1·4, *m*_2_ = 0·14 and *m*_1_ = 0·45, *m*_2_ = 0·15 respectively, so that the starting *R* remains around 1·3. In each situation, we keep *α* = 600 and distribute the vaccines (*a*) in proportion, (*b*) 50 percent to high-interaction and (*c*) 90 percent to high-interaction group and display the results in Table 1 below. For comparison, we also show the no-vaccine case.

**Table 1:** Outbreak duration and cumulative case count for three vaccine allocation strategies in three different social structurings. The symbol ‘k’ denotes thousand and ‘L’ hundred thousand.

| Vax \ Int. | 95—05 |  | 90—10 |  | 80—20 |  |
| --- | --- | --- | --- | --- | --- | --- |
|  | Days | Cases | Days | Cases | Days | Cases |
| **No vaccine** | 259 | 1.14L | 241 | 1.28L | 245 | 1.28L |
| **Proportional** | 216 | 61k | 214 | 71k | 221 | 66k |
| **50 percent** | 184 | 14k | 201 | 31k | 212 | 47k |
| **90 percent** | 181 | 8200 | 203 | 16k | 207 | 28k |

Thus, the greater the stratification in interaction, the more pronounced is the effect of preferentially vaccinating the high-interaction class. Although in Figs. 2-3 we can see an approximately 100-150 day interval before the case rates really become low, we **refrain** from advertising this as a prediction. This is because the durations were partly influenced by the parameters leading to the value of *R*; the 1000 cases/day in a population of 3,00,000 which we see in Fig. 2 might well overstress hospital capacity and force a lockdown, which will spin the trajectories out. Similarly, when the case rate decreases, reopening activities will start, which can again result in a surge and make the epidemic more protracted.

Now to come back to the question with which we started this Section, who gets the vax first ?

### ANSWER: After the healthcare workers are cleared out, it is the high-interaction people like shopkeepers and restaurant workers who should get the vaccine first

If you are a vegetable seller, bus conductor etc, that is obvious good news for you; otherwise you are at least assured that before your turn comes, the risk of your getting exposed to the virus at a store or in a bus will reduce considerably. Indeed, in Ithaca, New York State where one of us (SHAYAK) is located, the announcements from the Health Department regarding potential exposures at XXX store or YYY restaurant have become almost a daily menace after a recent climb in cases; vaccination in those places will ensure that we are more at peace visiting them before we get vaccinated ourselves.

### For policy makers

For the most effective utilization of the initially limited vaccine supplies, it is very important that we have a good idea of who is a high-interaction person and who is not. The best source for such a determination is the data obtained from contact tracing investigations. By analysing the secondary cases documented from each primary case, it should be possible to obtain a correlation between interaction rate and profession. Analysis of superspreading incidents should also cast significant light on this matter. Of course, incidents where an unmasked partygoer infected a whole crowd have to be ignored for this purpose.

We are also aware that there is considerable pressure on public health authorities to preferentially vaccinate the old and vulnerable people after the healthcare worker drive. While this reflects a noble sentiment, the maths might recommend a more balanced approach. We would expect most such people to belong to the low-interaction class and Table 1 shows an upto eightfold reduction in total cases arising from preferential vaccination of the high-interaction class. Therefore, while there is no doubt that among the low-interaction class the old and vulnerable should be vaccinated first, it might be more prudent to spend say half the doses on these people and the other half on high-interaction people, whatever their age. This might end up saving more lives overall than a vulnerable-first vaccination drive. A detailed analysis of the contact tracing data together with good estimates of the population structure etc will be necessary to arrive at an optimal strategy.

## §3 MOBILITY AND EFFECTIVITY

### QUESTION : I just got my vaccine. Can I take off my mask and party with others who have been vaccinated ?

This question is perhaps on everyone’s mind. On the one hand, we are by now fed up with masks and social restrictions and the shot seems to be the only get-out-of jail card. On the other hand, the shots are only partially effective. Suppose an attempted return to normal leads to a case explosion ?

To analyse this situation we start by assuming that when a high-interaction person receives the vaccine, his/her interaction rate remains the same but when a low-interaction person receives it, his/her interaction rate goes up at once. This is plausible because a shopkeeper will interact with an approximately fixed number of customers both during the pandemic and during normal life but work-from-home researchers like ourselves have a much more active social life during peacetime (some even more so than others). Thus, we expand the model (5-8) to accommodate three classes of populations – high-interaction, low-interaction unvaccinated and low-interaction vaccinated. Let *y, z*_1_ and *z*_2_ denote the case counts in the three respective classes and let *v* and *w* denote as before the vaccinated counts in the high- and low-interaction group. *N*_1_ and *N*_2_ remain the total high- and low-interaction populations. Now, we have three spreading rates – *m*_1_, that of the high-interaction class, *m*_2*a*_, that of the unvaccinated low-interaction class and *m*_2*b*_ (> *m*_2*a*_), that of the vaccinated low-interaction class.

The vaccination equations (5,6) remain almost unchanged. Vaccination of high-interaction people stops when *y*+*v* = *N*_1_ as before. Vaccination of low-interaction people now stops when unvaccinated cases plus vaccinees equal the total i.e. when *z*_1_+*w* = *N*_2_. This is an exact definition for the stopping condition of the vaccination drive; it corrects the error mentioned after (6) and shows us why the correction necessitates an increase in the system dimensions.

As for the case equations, for the high-interaction class, nothing changes from (7) except for the number of at large cases, which must now include all case categories. We have

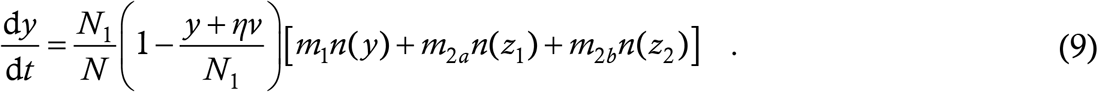

The total number of at large cases stays the same in the two remaining case equations as well. Among the low-interaction class, at any instant, the number of unvaccinated people is *N*_2_ − *w* and the number of vaccinated people is *w*. Thus, for the *z*_1_-equation, the probability that a random target belongs to the unvaccinated low-interaction class is (*N*_2_ − *w*)/*N*, which replaces the *N*_2_/*N* of (8). Then, the number of susceptible people among this class is everyone save those who have had the infection already i.e. it is *N*_2_ − *w* − *z*_1_, and the probability of susceptibility is that divided by *N*_2_ − *w*. Putting this together yields

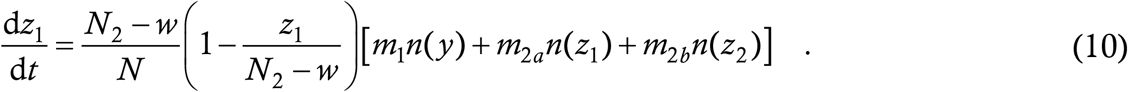

For the vaccinated low-interaction class, the first term on the above RHS gets replaced by *w*/*N*. By definition of vaccine effectivity, a fraction 1−*η* of all the vaccinees are susceptible. Among these however, the cases *z*_2_ will have to be excluded since they have contracted the infection already. Thus, the total number of susceptible people of this class is (1−*η*)*w* − *z*_2_, and the susceptibility probability is that divided by *w*, which is (1−*η*) − *z*_2_/*w*. Putting all this together, we get

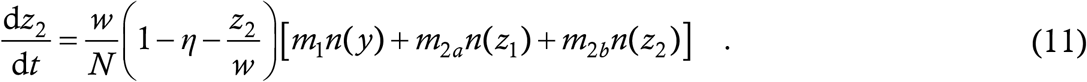

Equations (5,6,9-11) make up the system which we shall have to simulate.

We use the 90—10 social structuring from the previous Section. We also adopt the values *m*_1_ = 0·75 and *m*_2*a*_ = 0·15, which we recall contributes an average *R*_0_ of approximately 1·3 before the vaccination starts. For *m*_2*b*_, we note that the calculated *R*_0_ for COVID-19 in the early phases of spread before any interventions were instituted [4] is approximately 3. When the unvaccinated low-interaction class disappears, the value of 0·47 for *m*_2*b*_ would result in an average *R*_0_ close to 3. Hence we take this value here. We take *α* = 600 and *β* = 0·5, giving high but not overwhelming priority to the high-interaction class. For the display Figure below, we take the effectivity value *η* = 0·90. Blue line is high-interaction cases, green line low-interaction unvaccinated cases and red line low-interaction vaccinated cases; grey, cyan and yellow bars are the respective epi-curves.

Having trouble seeing the yellow bars ? They exist. Zoom the picture a thousandfold and you’ll find them just above the ground-hugging red line. That’s how many cases crop up among the vaccinated people after they return to normal life. More formally, the epidemic ends at 197 days with 1,15,000 vaccine doses being required. The total case count is 27,123 of which 158 are in the vaccine group. With the effectivity reduced to 0·6, the curves are below.

This time, the yellow bars are visible but still quite small. The epidemic formally ends at 316 days with 65,000 cases of which 2065 are in the vaccine group. Further reducing the effectivity does result in trouble however. At *η* = 0·5, the epidemic runs for 948 days with total just above one lakh cases; almost 20 percent of these are in the vaccine group. In the limit that the vaccine is a mere placebo, about 30 percent of the total 2,28,000 cases occur in the “vaccine” group. The fraction is not higher because a substantial fraction of the cases occurs even before the majority of people receive the dummy shots and become more mobile. Thus, we can say that for these parameter values, there is a sharp transition between an effectivity of 50 and 60 percent. In the high effectivity regions however, there is no cause for concern. Once again, in this situation, the value of *β* (allocation preference) plays a very important role. In the 90 percent effectivity case for example, *β* = 0·1 increases the final case count from 150 to just above 1000 while *β* = 0·9 reduces it to just 20. We are not seeing any **qualitative** change however with a change in *β*.

In Fig. 4 we can see a duration of about 120 days until the epi-curves become minuscule. Unlike in the last Section, this time we have greater confidence in the duration as a reasonable predictor of reality. This is because a reopening plan is built into this analysis – at the end of the epidemic, society is in a full-mobility state rather than a constrained mobility state. The approximate duration comes from a heuristic consideration : the peak should come when *R* = 1. Let us ignore the small contribution of vaccinees to *R*; the others contribute a starting value of 1·3 which reduces to unity when about 1/4 of the people have been vaccinated. With 500 days to vaccinate everyone, about 125 days suffices to achieve this vaccination level and hence the peak. As it turns out, the maths generate a more satisfying result than the heuristics. Returning to the starting question, should a vaccine confer immunity against social restrictions ?

**Figure 3:**
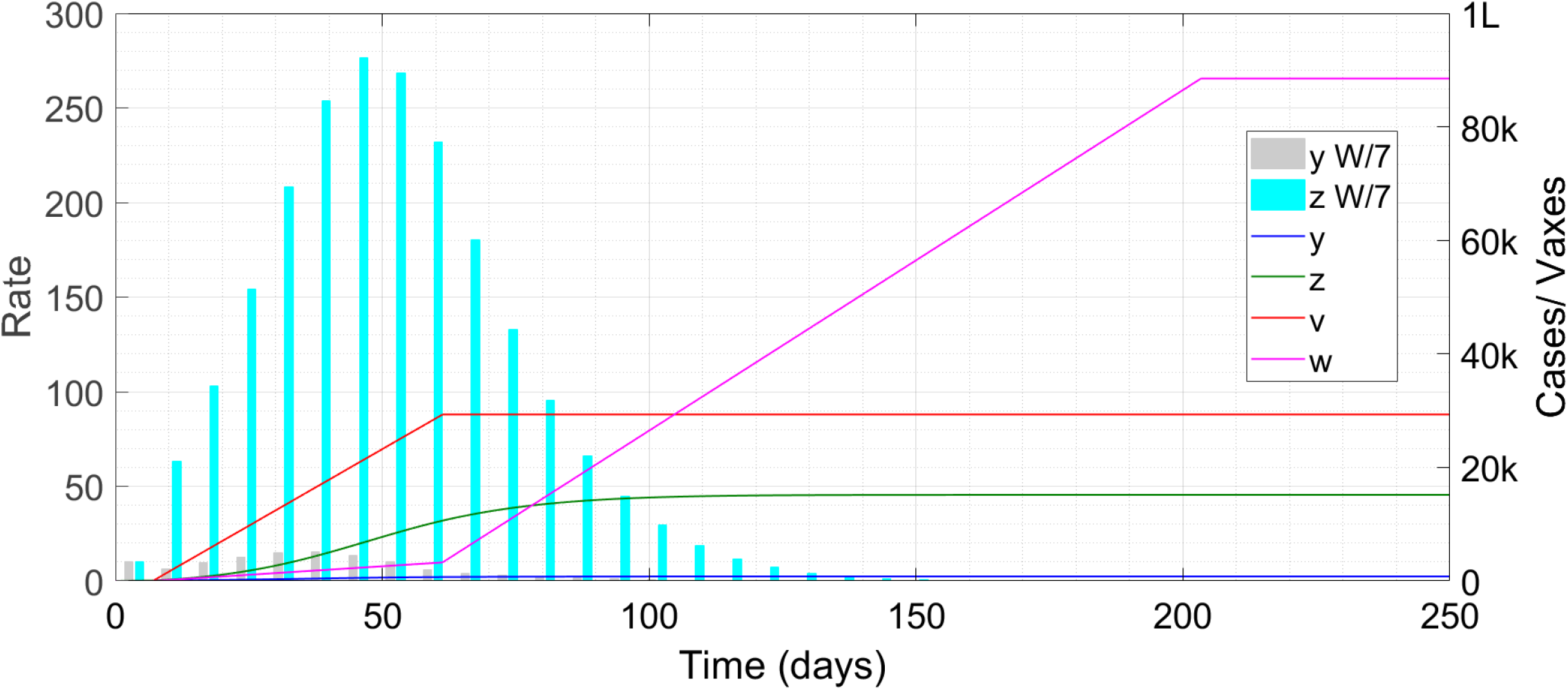
Time trace of the epidemic with vaccination. The symbol ‘k’ denotes thousand, ‘L’ hundred thousand and ‘W/7’ weekly cases scaled down by a factor of seven.

**Figure 4:**
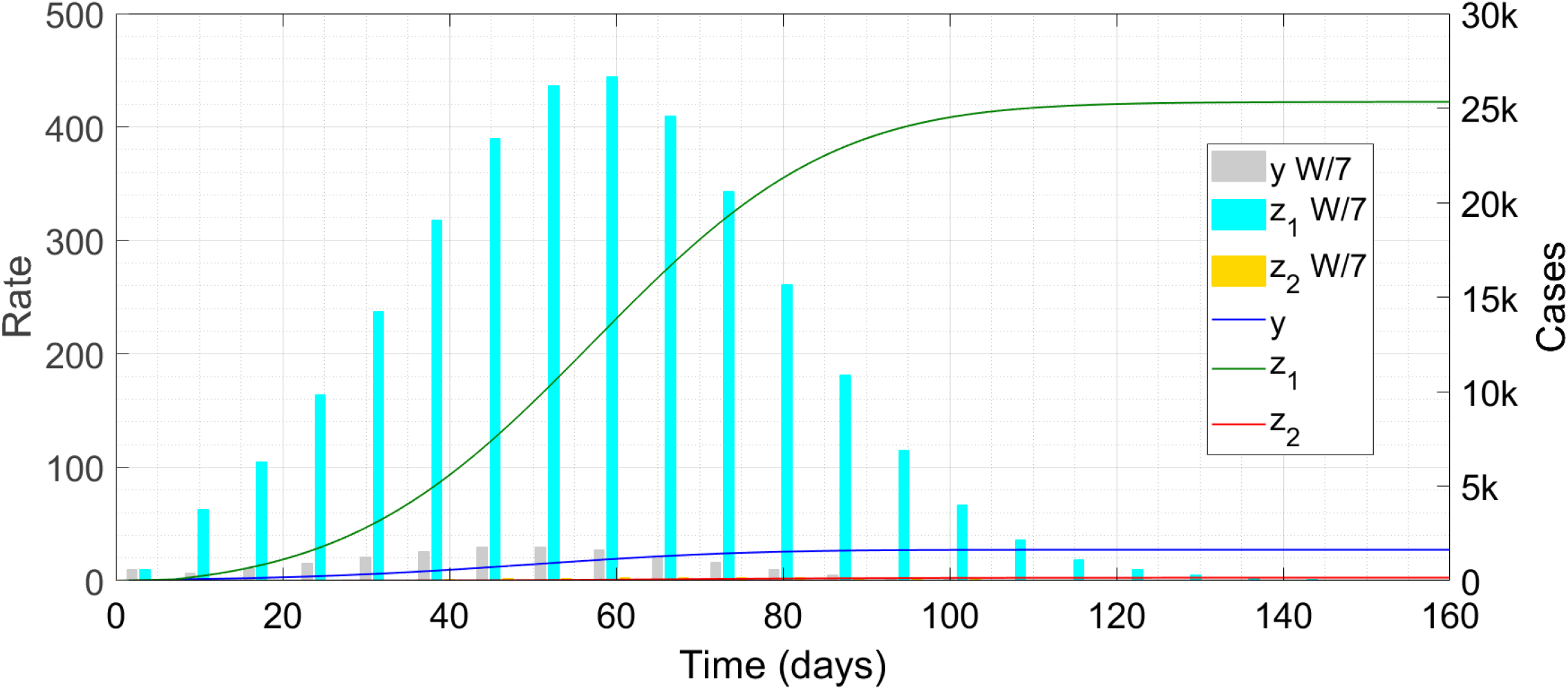
Time trace of the epidemic with increased mobility for vaccinees. The symbol ‘k’ denotes thousand and ‘W/7’ weekly cases scaled down by a factor of seven.

### ANSWER: Cognizant of what we are saying, we declare yes

The data released so far does seem to indicate that *η* = 0·9 is closer to reality than *η* = 0·6, and with this kind of effectivity there is hardly any contribution to cases if the vaccinated people take off their masks and get about their normal lives. One way of understanding this is in terms of *R*. The unconstrained *R* (i.e. *R*_0_) of COVID-19 is approximately 3. This means that, when people lead normal lives, each case spreads the disease to approximately 3 more people. If the vaccine is 2/3 effective, then 2/3 of the vaccinees will be immunized; if the vaccinees lead a normal life then one case will spread to only one more person. Make the effectivity higher and this becomes less than one, and the epidemic dies down. Still, you are not convinced – “A 90 percent effective vaccine means 10 percent chance of contraction. That’s low all right but not exactly zero; are you sure I can take off my mask and have fun ?” We get your point, and agree that 10 percent is low but not minuscule. So, if you are vaccinated, the probability of your contracting the virus **given an exposure** reduces by a factor of 10. But, contraction is a two-person process; first, the person you are interacting with has to expose you and then you have to catch it. If you socialize only with other vaccinees, then the probability that the other person has it also goes down by a factor of 10, so the probability of your getting exposed reduces by 10 and the probability of your contracting it then goes down by 100. That is a far more reassuring reduction. Since transmission is interactive, the contraction probability is **quadratic** rather than linear in the vaccine failure probability.

### For policy makers

This is probably a surprising result, so we have provided two intuitive justifications apart from the math. Intervention fatigue has already taken root in society, and attempting to force restrictions on vaccinees will be a thankless task. Fortunately, our analysis seems to indicate that such enforcement is not required. Relaxation of social restrictions for vaccinees means that there has to be a system for authorities to identify them, such as an I-card given to every vaccinee at the time of receiving the second dose. For selective restrictions in public places, it will also help if vaccinees can be identified easily by sight – this might be facilitated by distributing apparel or badges which advertise a person’s vaccinated status. Such identifying mechanisms might also be used to relax domestic and international travel restrictions for vaccinees. The authorities might be hesitant to take masks off high-interaction people like shopkeepers even after they have been vaccinated – suppose they turn superspreader ? As a compromise, such professionals might initially be required to have masks on during work hours but take them off after hours.

Removing restrictions for vaccinees is possibly the easiest and safest way of returning to normal from the present situation. Blanket policies applicable to everyone might meet with more resistance all across the board, and might also result in case surges when restrictions are collectively lifted. With this strategy, a collective relaxation can of course be announced when case prevalence has really decreased everywhere. The selective relaxations also mean that people who get the shots later will be envious of peers who have got it earlier. To minimize such heterogeneity, entities such as universities and small employers which arrange for vaccination of their constituent members might wait to start the drive until they have acquired enough doses to vaccinate all the members at one go. Of course, employees who are old or have comorbid conditions cannot be involved in this waiting game and must be immunized as soon as a batch of shots is available. Finally, if a particular vaccine is such that the authorities feel no confidence to relax restrictions for the vaccinees, for example due to hospitalizations and/or deaths, then the authorization of that vaccine probably needs a second look. Any vaccine whose recipients cannot be cleared for normal life is probably not worthy of manufacture or distribution.

## §4 EFFECTIVITY VS EFFICACY

### QUESTION : They tell me the vaccine efficacy is 90 percent based on indirect observations. Does it mean that the vaccine confers immunity with 90 percent probability or some other probability ?

We have already introduced the difference between efficacy *E* and effectivity *η* in Sections 0-1. Here we see how the two are related. Following the last Section, we now augment the basic model (5-8) into four classes – unvaccinated and vaccinated high- and low-interaction people. Let *y*_1_, *y*_2_, *z*_1_ and *z*_2_ denote the case counts among these classes, and going with the most general situation, let there be four *m*’s, *m*_1*a*_, *m*_1*b*_, *m*_2*a*_, *m*_2*b*_. Then, repeating the logic of the last Section leads to the case equations

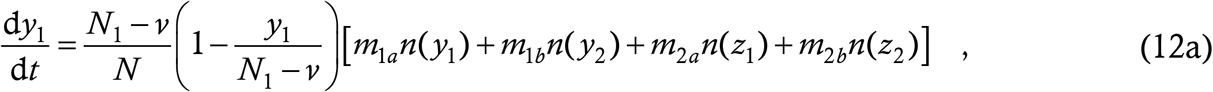

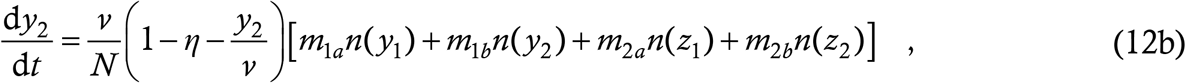

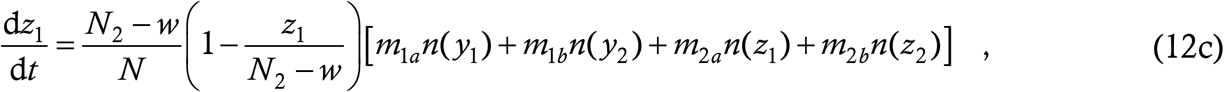

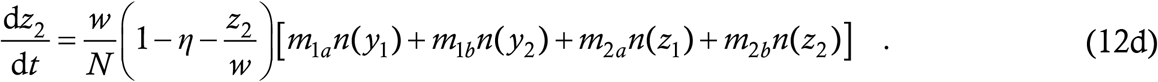

The vaccination equations remain the same as (5,6) except that *y* and *z* in those equations must be replaced by *y*_1_ and *z*_1_.

While the efficacy so far has been defined in terms of trials, with equal numbers of participants being recruited in the vaccine and placebo groups, we now need a definition which extends this from “the lab” to “the field” with the epidemic and the vax drives running in earnest. In this situation, we define the efficacy in terms of the case rate normalized to the population in the unvaccinated and vaccinated groups. This makes the efficacy a function of time. The mathematical definition we use is

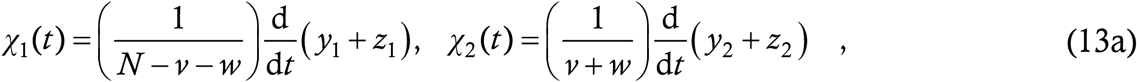

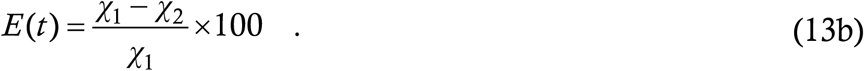

The use of the case rate rather than the cumulative count ensures that errors arising from a high initial rate in the unvaccinated group do not persist with time.

We now plot the efficacy (13) as a function of time for the simulation run of Fig. 3 (with *m*_1*a*_ = *m*_1*b*_ = 0·75, *m*_2*a*_ = *m*_2*b*_ = 0·15), where effectivity is taken as 80 percent.

We can see that the efficacy works out to nearly 80 percent as well during the entire outbreak. As another test case, we rerun the situations of §3, using *m*_2*b*_ = 0·47 for the vaccinated low-interaction people. With *η* = 0·9 (situation of Fig. 4), the efficacy works out to 90 percent throughout the epidemic evolution, just as in Fig. 6 above; with *η* = 0·6 (situation of Fig. 5), we see the following.

**Figure 5:**
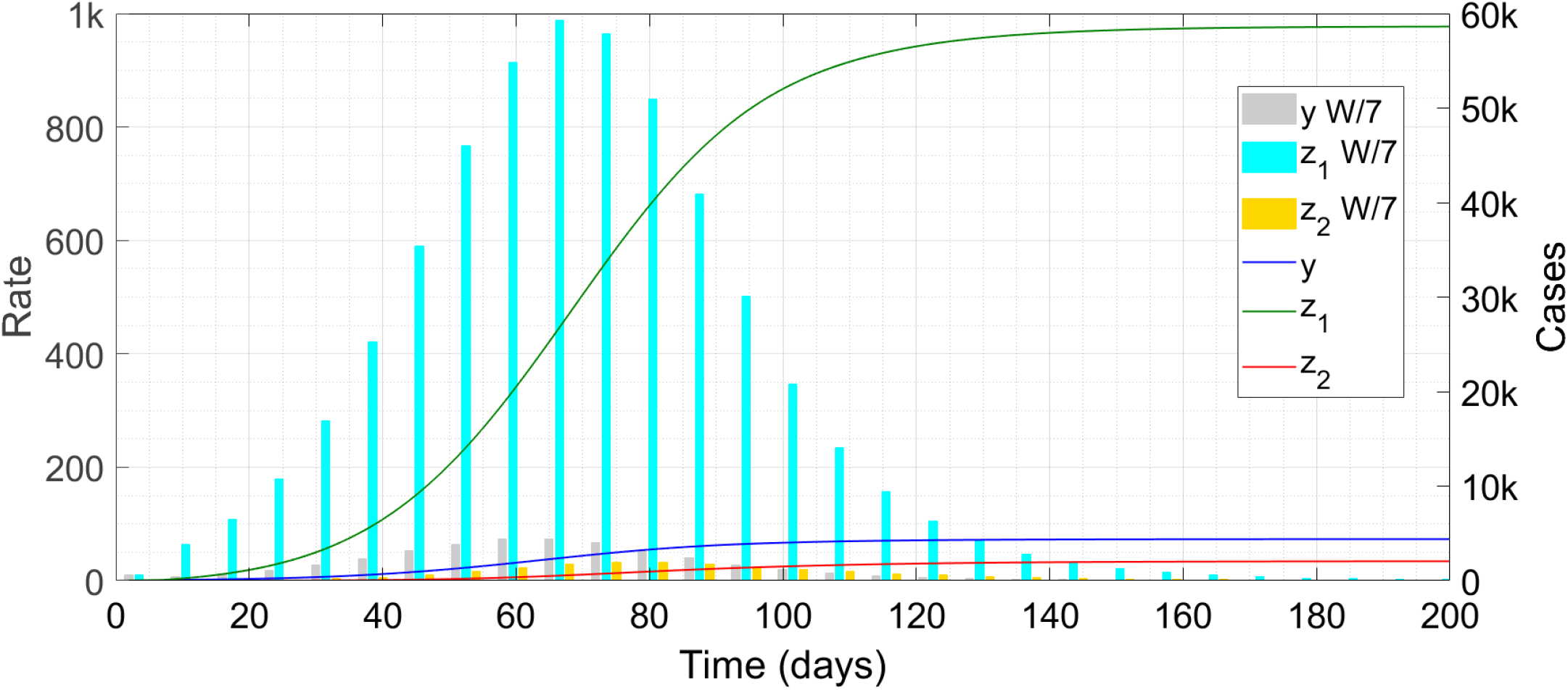
Same as Fig. 4 but with a less effective vaccine. The symbol ‘k’ denotes thousand and ‘W/7’ weekly cases scaled down by a factor of seven.

**Figure 6:**
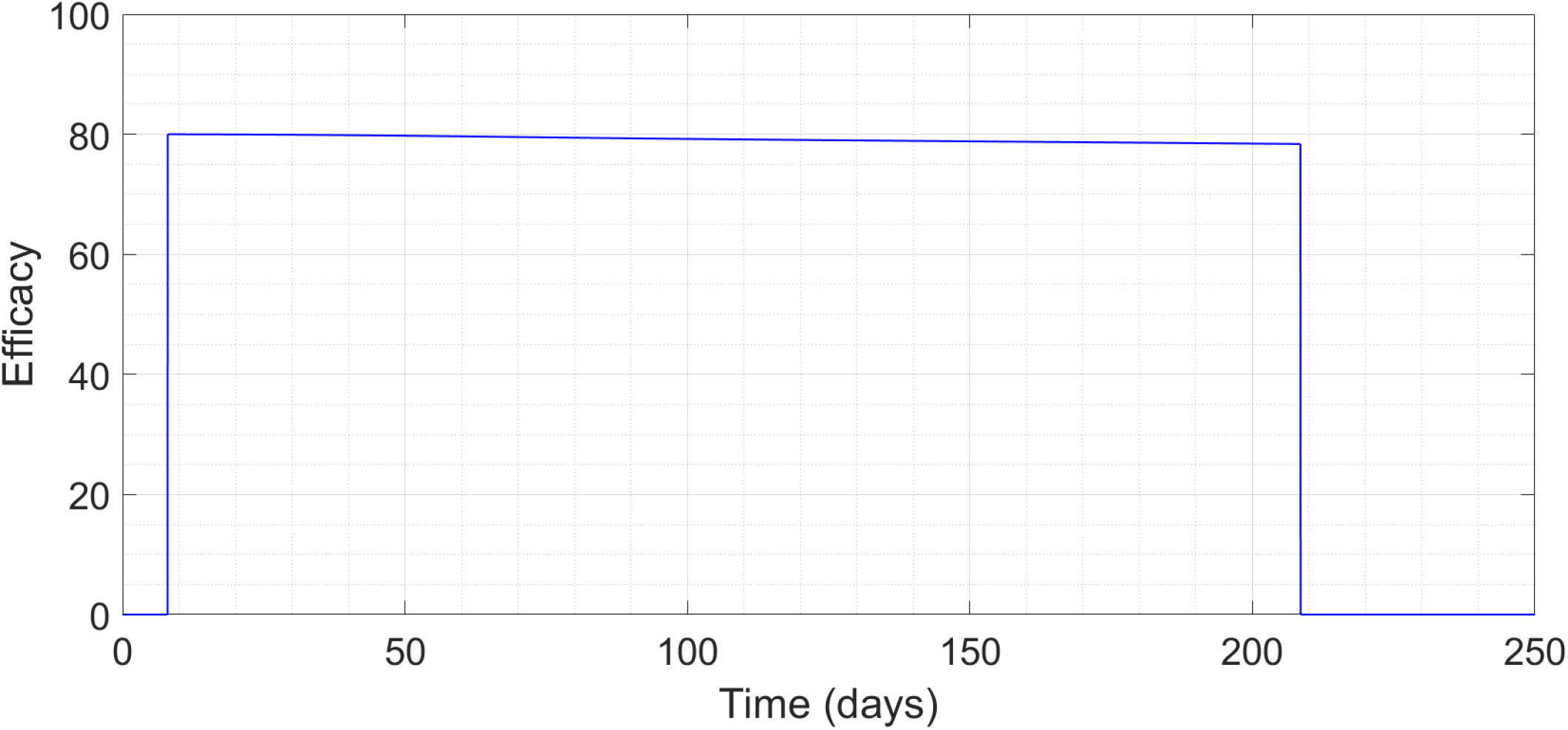
Efficacy as a function of time for a vaccine with effectivity 80 percent.

This time, the efficacy starts from the effectivity but decreases slowly throughout the epidemic evolution. Note however that it remains quite close to the value 0·6 during the first 150 days, when the case rates are significant. From this run, we also find that the total case count at the end of the run is 63,000, of which 1984 are in the vaccine group. These numbers are slightly lower than the 65,000 and 2065 obtained in §3, which involves the approximation of §1 in the *y*-equation. Consistent with our reasoning, the approximation has caused the reported case counts to exceed their true values.

So are efficacy and effectivity the same ?

### ANSWER : Yes, it does appear that if they tell you the vaccine is 90 percent efficacious, then the probability of your being immunized by it will be 90 percent

Of course there can be a difference of a few percentage points depending on how the epidemic evolves etc but it is very very unlikely that a vaccine of efficacy 90 will have a true effectivity of 60.

### For policy makers

Equation (13) shows a reasonable extension of the definition of efficacy from a vaccine trial to a real world scenario. It also indicates that the efficacy defined indirectly in this manner is very close to the true effectivity which would have been obtained from a challenge trial. The simulations seem to suggest that if anything the efficacy underestimates the effectivity. Another source of error will come from the limitations on our knowledge of the value of *N* – we must go with the entire population of say a city, but the true value will probably be a fraction of it. Using a higher *N* in (13a) reduces *χ*_1_ below its true value and hence reduces the calculated efficacy. Thus it is unlikely that an efficacy calculated in this way shall yield a value higher than the true effectivity and inspire false security among authorities and vaccinees.

## §5 TEMPORARY IMMUNITY

### QUESTION : Will there be waves of corona outbreak even after the vaccines are widespread ? Will I need to go to the vaccine clinic every few months ?

With any infectious disease, a very important question is how long does one bout of infection render a person insusceptible. We have carried out an extensive analysis of this situation in a prior work [2] which obviously doesn’t include vaccination. There we have shown that if immunity lasts for a duration *τ*_0_, then the spreading characteristics are determined primarily by whether the evolution with permanent immunity lasts shorter or longer than *τ*_0_ (this is not an exact condition but a very good approximation). In the former case, the disease progresses to a halt after a single wave while in the latter case it can run on forever in wave after wave. Rapid progression of the disease is achieved by a higher *R*; thus, a more relaxed intervention level, although producing higher case counts initially, can be more beneficial on the long run. This is a qualitatively intuitive result – if the outbreak is over before the first patient has lost immunity, then surely it does not matter how long immunity lasts. For more details, please consult Ref. [2].

This Reference also contains a review of a large number of Literature items which have appeared on the subject of COVID-19 immunity. To summarize this review, it does seem that immunity lasts for at least eight to nine months following infection in the overwhelming majority of patients – among the crores of cases discovered since March, so far there have been only nine reinfections confirmed through phylogenetic analysis. Robust antibody response has been detected in most patients at upto 5 months following infection; significant concentrations of T- and B-cells have been detected at 6 months even among patients with low or negligible antibody titre levels. We refrain from citing the original antibody references again but refer to Ref. [2] for the relevant items, which are about 30 in number. What is totally unknown as yet is the duration of immunity conferred by the vaccine – that shall become apparent only as the disease evolves. Hence we shall have to work here with some assumed values.

With the vaccination drive in place, we would again expect one-shot and multiwave solutions with an approximate condition separating the two being that everyone should get immunized (via either the disease or the vaccine) within the immunity period. Thus, intuitively, the vaccination drive (and/or the outbreak itself) must progress at a fast enough pace for the disease to be eliminated in time. How fast is fast can no longer be obtained from intuition; for that we must run the mathematical model.

To extend the model (5-8), we proceed as in Ref. [2]. Let both the disease and the vaccine confer immunity for *τ*_0_ days following infection/vaccination, after which the case/vaccinee becomes completely susceptible to the disease again (this corresponds to the **simple immune response** of Ref. [2]). The only term in the structure (0) which depends on the immune response is the susceptibility probability. In the equivalent of (7), the pool of insusceptible people at any instant comprises everyone who has contracted the disease within the last *τ*_0_ days and a fraction *η* of all the people who have been vaccinated within the last *τ*_0_ days. This is mathematically expressed as *y* − *y*(*t* − *τ*_0_) + *η*(*v* − *v*(*t* − *τ*_0_)), and, carrying through the remaining steps we get

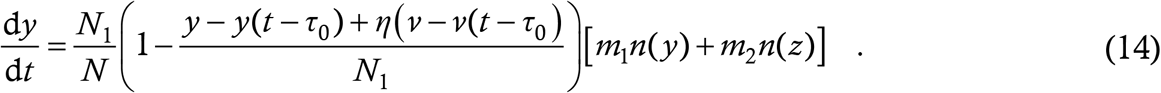

Analogously, the *z*-equation reads as

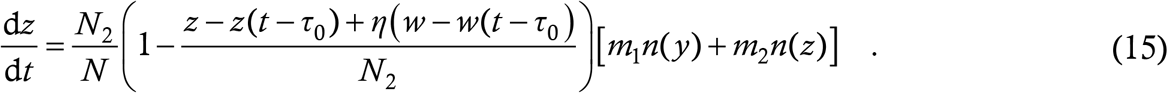

As for the vaccination equations, if immunity is temporary then the vaccination drive must go on so long as the disease exists. For our analysis however, it is sufficient to consider one round of vaccination, described by the equations (5,6). If the disease stops within that round itself, then we can declare elimination, while if there is a resurgence after the drive is over then we need to extend the drive. Hence, we keep (5,6) unchanged and club them with (14,15) to form the model.

In Ref. [2] there are two scenarios where we have found multiwave trajectories with temporary immunity – one is where parameter values remain constant so as to generate a somewhat-above-unity *R*_0_, and the other is where parameter values change with time to generate an increase in *R* midway through the outbreak. We have labelled these as “City B” and “City G” solutions respectively. In other scenarios, the solutions have remained the same with temporary and permanent immunity. With the vaccination drive, we expect special solutions in similar regions of the parameter space, and run the simulations accordingly. For all simulations, we choose the 90—10 social structuring and take the vaccine effectivity to be 80 percent.

For the first run, we take *τ*_0_ = 200 days, as in Ref. [2]. This is a gross underestimate for the disease; it might hold true for the vaccine especially if these first-generation vaccines are not optimal at eliciting a durable immune response. Nevertheless, 200 days should be treated as something of a worst case scenario. Firstly, we note that the runs of Figs. 2-4, which feature highly effective vaccines, all terminate within or just above 200 days. These don’t change if the immunity period is taken to be this value. A City B-type solution can be generated if we use the parameter values *m*_1_ = 0·70, *m*_2_ = 0·14. We first display the solution function without vaccination.

We have shown here the first two of infinitely many waves of outbreak – the picture is identical to Fig. 3 of Ref. [2]. In this situation, with *β* = 0·1 (no preference to high-interaction class), the cutoff between one-shot elimination and perpetuation occurs at *α* between 370 and 380; we show the two cases in the Figures that follow.

Thus, a very modest rate of vaccination (at 380/day, it will take 2·5 years to vaccinate everyone) has sufficed to eliminate the disease despite the short-term immunity. Once again, it pays to vaccinate the high-interaction people first. Instead of *β* = 0·1, if we use the value *β* = 0·9, then the cutoff *α* comes down to between 210 and 220 – a 40 percent reduction from the previous case.

Of course, in the runs we considered so far, the elimination was facilitated by public health measures – low *R* throughout means continued observance of restrictions which is unrealistic in practice. Hence we now consider a City G-type scenario where reopening starts once the epidemic comes under control. For this set of runs, we increase the immunity period *τ*_0_ to 350 days, which is certainly more realistic than 200 for the disease itself. We keep *m*_1_ = 0·75 throughout, start off *m*_2_ with the value 0·15 and increase it linearly to 0·45 during the period from 200 to 350 days.

With no vaccine, there are two waves of outbreak as in the Figure below.

The second wave here is the result of the reopening and not of the limited immunity, as in Fig. 4 of Ref. [2]. This wave infects so many people that the epidemic itself becomes dead after it is over. If vaccination is introduced at rate *α* = 500 and *β* = 0·1 then we get the following.

“Can’t be” is your initial reaction – four waves instead of two ! But that’s how it is. The first wave and the second wave (reopening-induced) are lower and broader on account of the presence of the vaccine. This allows the epidemic to simmer on after the second wave, and when sufficient people from the first two waves have lost immunity, there is the third wave. This wave occurs shortly after the vaccination drive has stopped, so it is still a somewhat muted explosion which does not blow up the virus itself. Rather, the embers keep glowing and when the vaccinees also enter the susceptible pool, they ignite a conflagration. Only thereafter is the scourge over. Of course, we must remember that the social restrictions as well as vaccination drive remained off during the latter two waves. In practice, the beginnings of the third wave would necessitate the whole rigmarole all over again. This time, we need *α* of about 720 to terminate the epidemic in one go – the case count at the end is about 9000. This amounts to vaccination of the entire population in a little more than one year and might be a logistical challenge. Once again however, we can gain hugely by prioritizing the high-interaction class. At *β* = 0·9, the cutoff *α* reduces by almost half to 410. As an aside, note that the second wave in Fig. 12, which does not depend on temporary immunity, is an example of how a collective reopening plan can backfire. Such a phenomenon cannot occur with the selective reopening plan of §3, which demonstrates the superiority of the latter plan. Bottom line, do you need periodic vaccination ?

**Figure 7:**
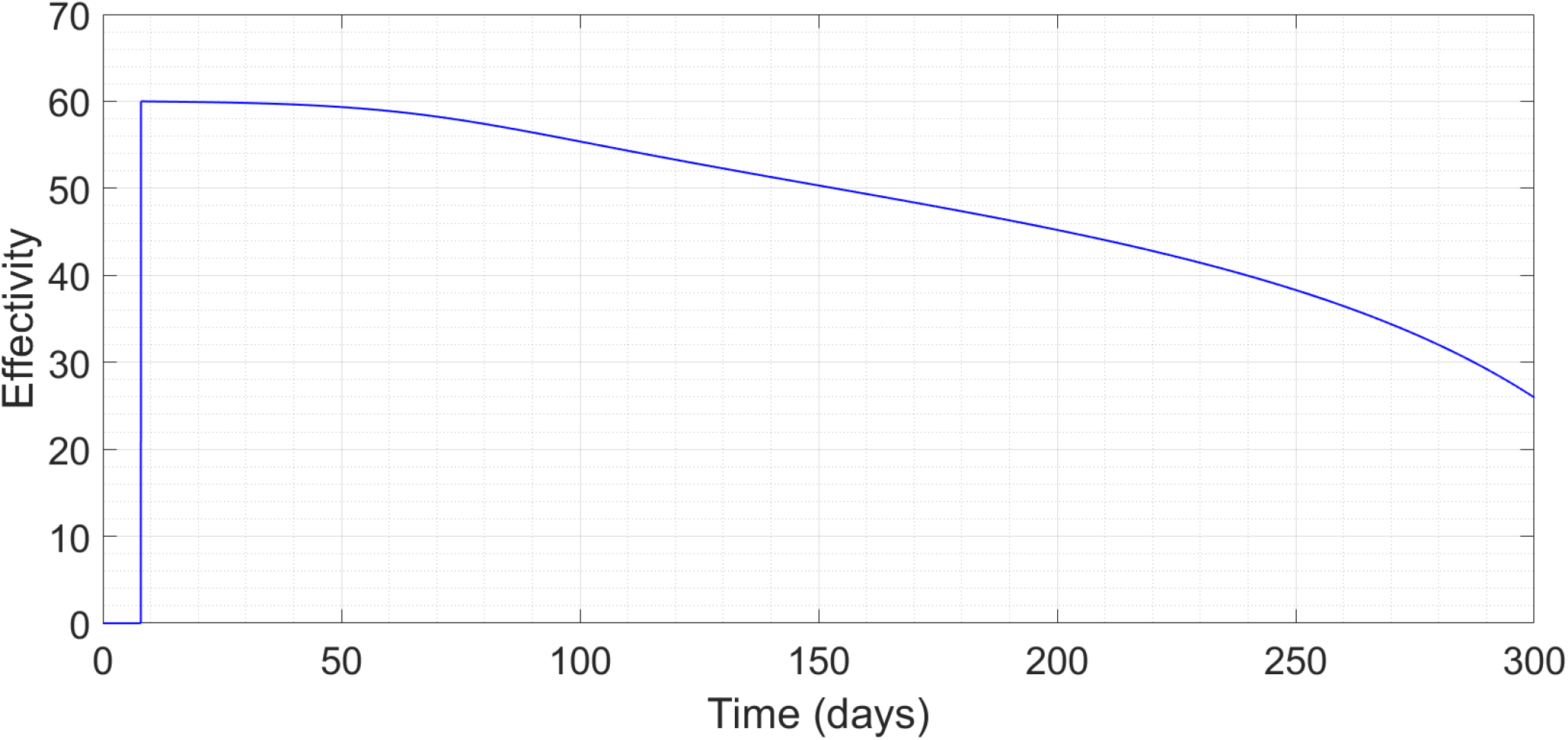
Efficacy as a function of time for a vaccine with effectivity 60 percent.

**Figure 8:**
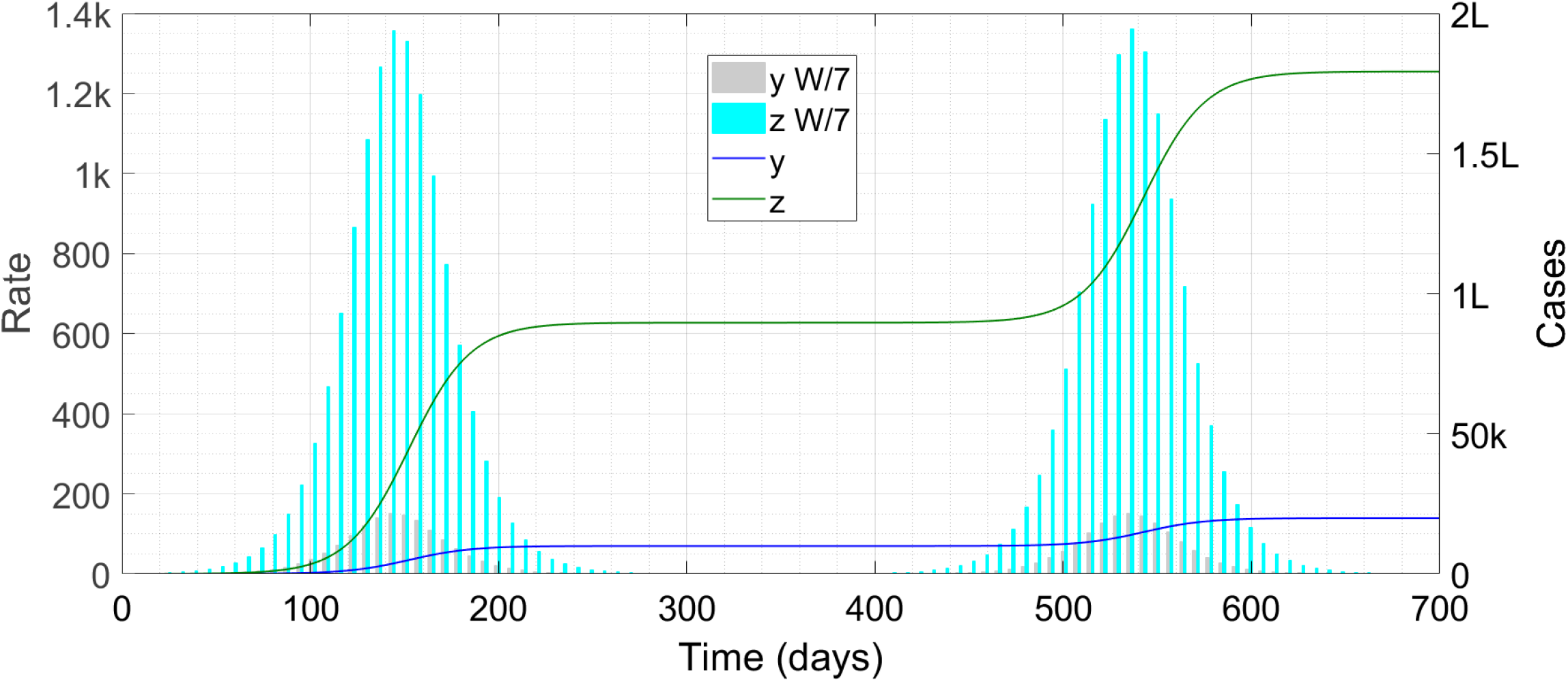
Time trace of the epidemic with temporary immunity and no vaccines. The symbol ‘k’ denotes thousand, ‘L’ hundred thousand and ‘W/7’ weekly cases scaled down by a factor of seven.

**Figure 9:**
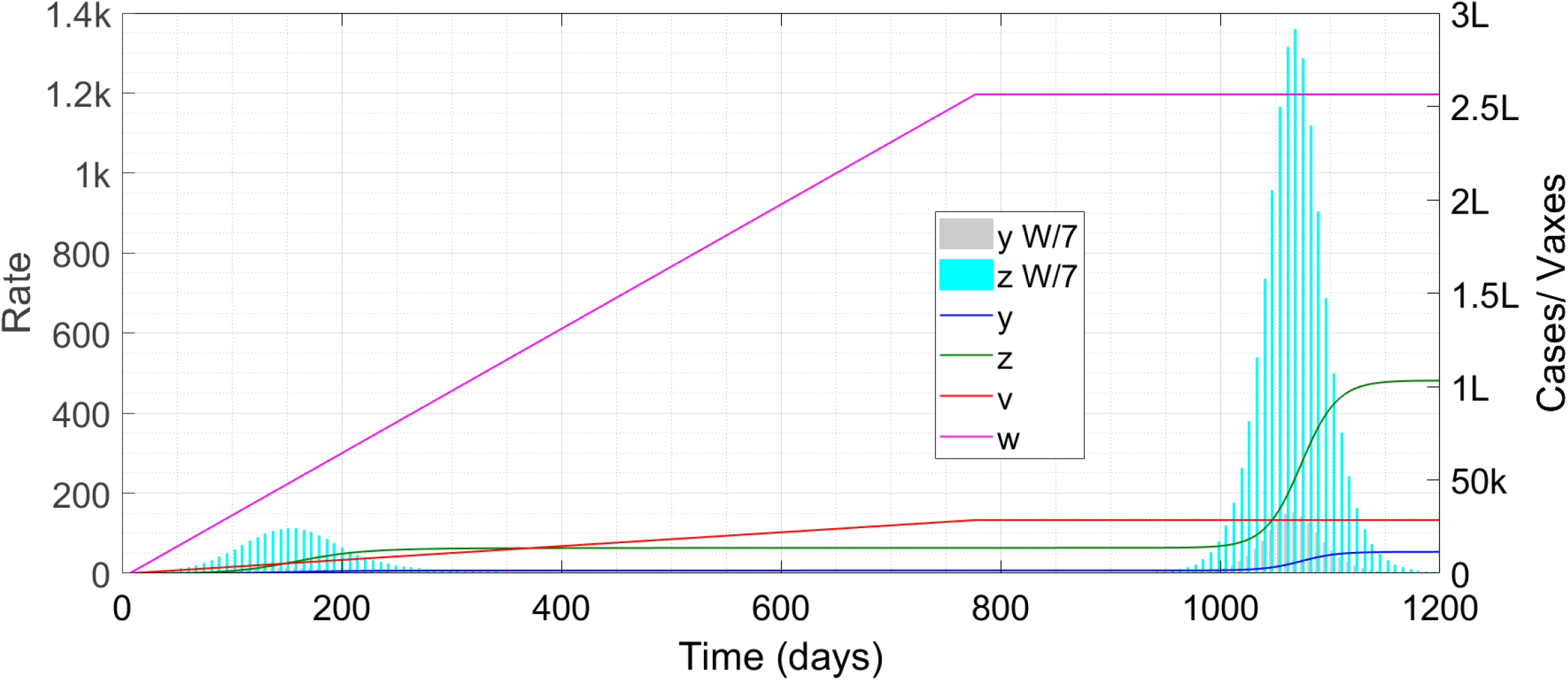
Time trace of the epidemic with temporary immunity and vaccination rate just below the elimination cutoff. The symbol ‘k’ denotes thousand, ‘L’ hundred thousand and ‘W/7’ weekly cases scaled down by a factor of seven.

**Figure 10:**
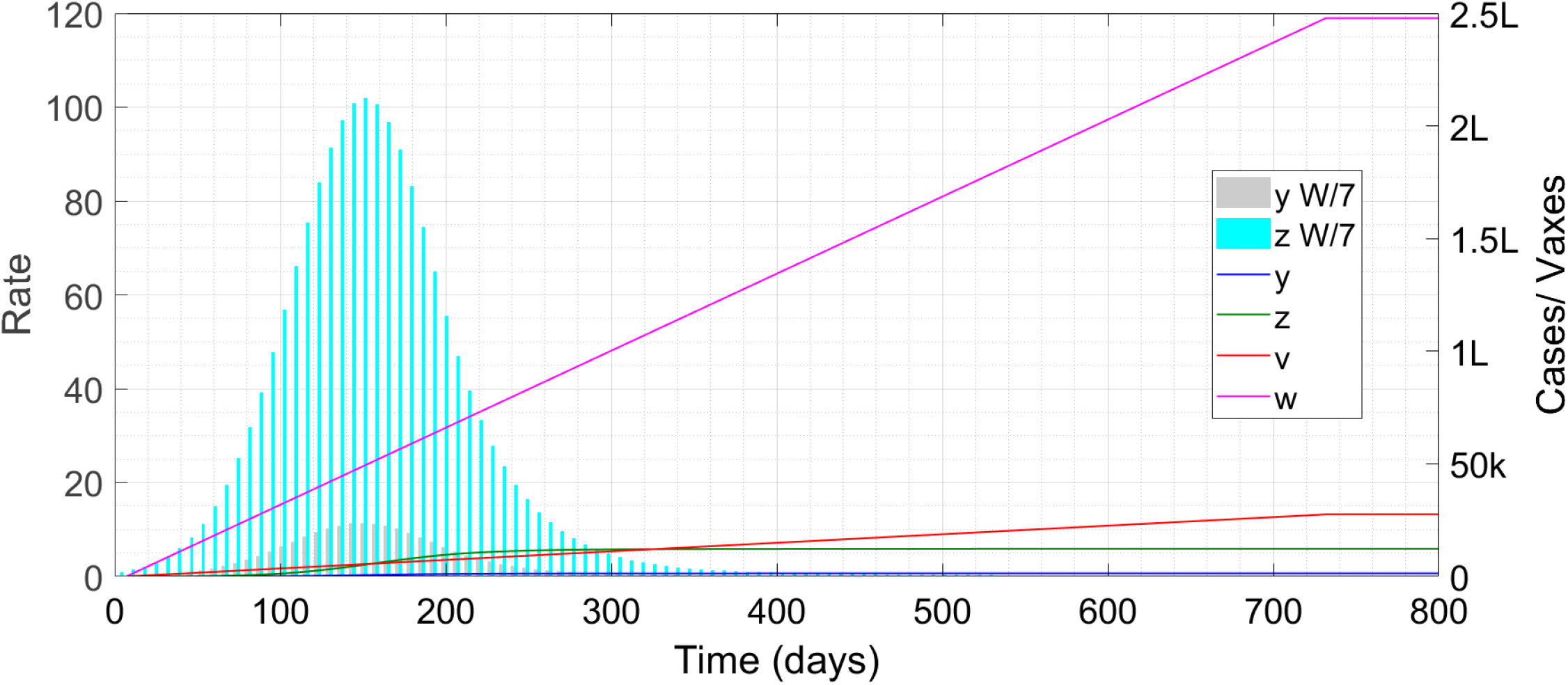
Time trace of the epidemic with temporary immunity and vaccination rate just above the elimination cutoff. The symbol ‘k’ denotes thousand, ‘L’ hundred thousand and ‘W/7’ weekly cases scaled down by a factor of seven.

**Figure 11:**
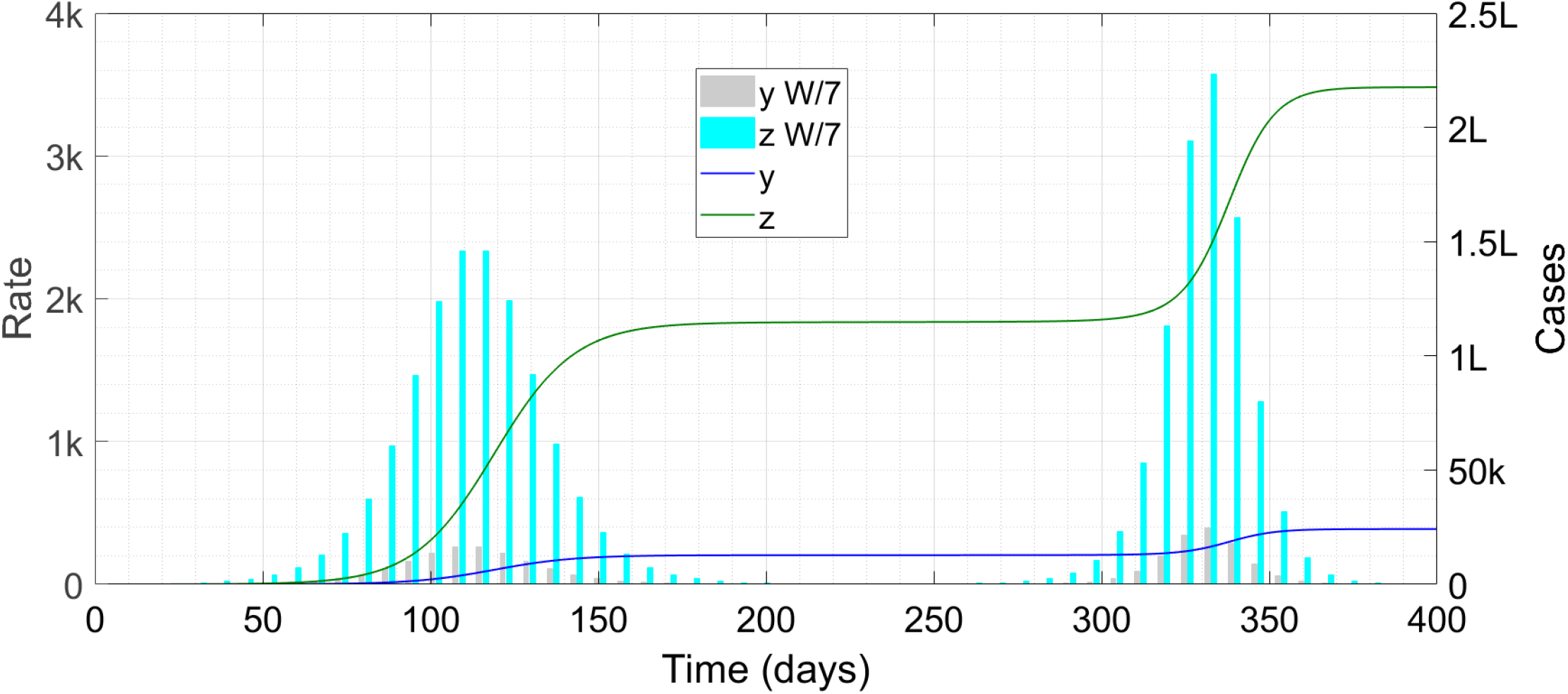
Time trace of the epidemic with temporary immunity, progressive reopening and no vaccine. The symbol ‘k’ denotes thousand, ‘L’ hundred thousand and ‘W/7’ weekly cases scaled down by a factor of seven.

**Figure 12:**
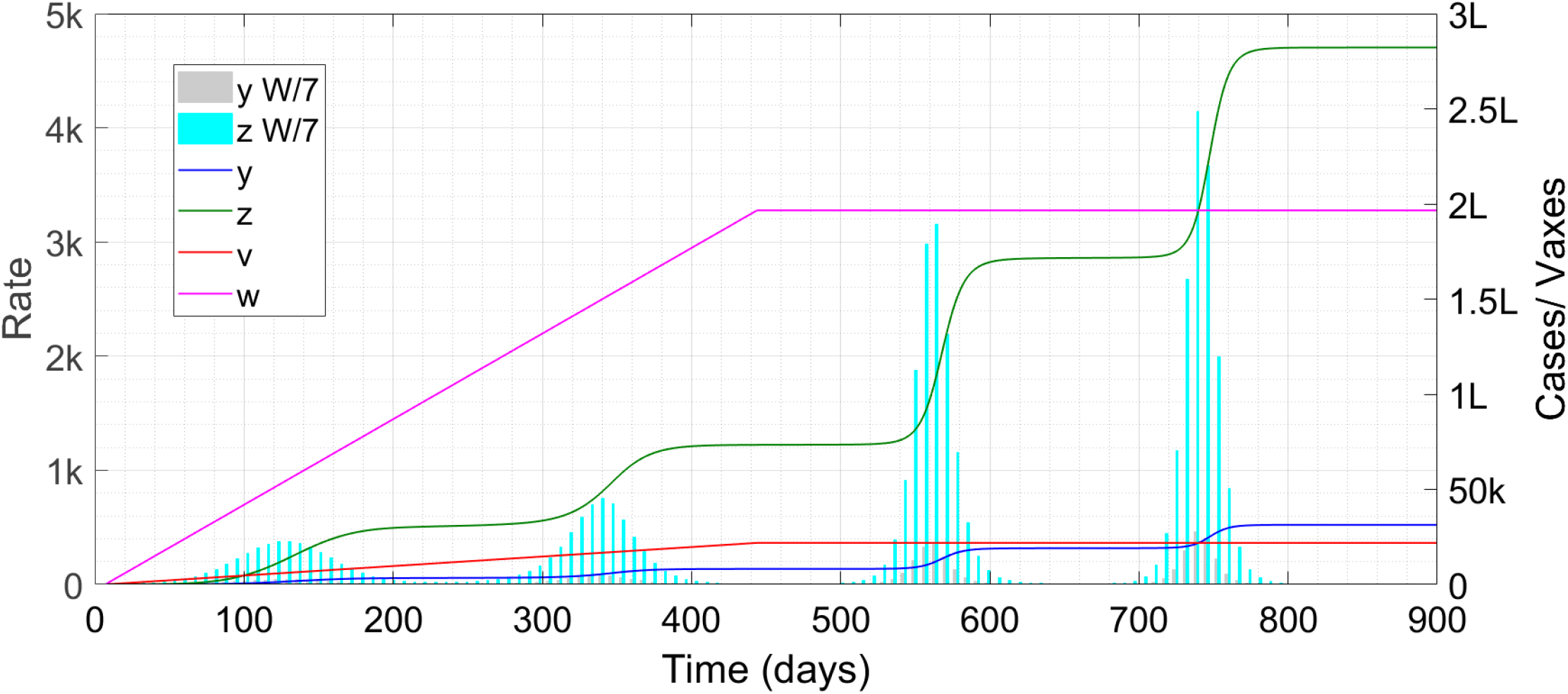
Time trace of the epidemic with temporary immunity, progressive reopening and inadequate vaccination rate. The symbol ‘k’ denotes thousand, ‘L’ hundred thousand and ‘W/7’ weekly cases scaled down by a factor of seven.

### ANSWER : The best we can say right now is that it depends

So long as the disease is in the air, you will have to repeat your shots as often as needed to stay immunized. With widespread vaccination however, elimination of the disease is definitely possible even if immunity is temporary. Among the situations we considered, only in the worst case (situation of Fig. 11) did we actually need to vaccinate close to everyone within the immunity period – in the other scenarios, we got by with a significantly lower cutoff vaccination rate. Given the havoc which the disease has caused, the authorities will certainly be desperate to drive it to oblivion, and will pull out all stops to ensure that it happens. Vaccine-driven eradication of infectious diseases has occurred in the past, with smallpox (variola major/minor) being the most notable example. In this case also, the immunity conferred by the vaccine was not lifelong but that did not prevent the disease from eventually disappearing forever.

“I have to take the flu vaccine every year,” will be a potential reaction. That is because the influenza virus mutates rapidly and the vaccine administered in one year is ineffective against the viruses which crop up next year. Coronaviruses in general mutate much more slowly, and the COVID-19 virus seems to be no exception. If the immune response generated by infection or vaccine is long-lived, then the mutation rate can also act as the basis for defining *τ*_0_ – the time after which the virus has mutated enough to make the existing antibodies useless or only partially effective. If the mutation rate can be established, then vaccination drives will have to try beating that rate and stamping the disease out. Another route to elimination can open up if it turns out that the vaccine provides durable severity-reducing immunity even if short-term sterilizing immunity (the **complex immune response** of Ref. [2]). In that case, once it is established that vaccinees will only suffer a mild disease, a natural outbreak among these people can result in the disappearance of the disease.

In summary, there are far too many unknowns yet, and elimination of the virus cannot be guaranteed. All that our simulations show is that it is by no means an impossible outcome. Even if not completely eradicated, it might still be possible to confine the virus to sporadic outbreaks in extremely localized regions, as happens with the Ebola virus. We do expect that research into vaccine development will continue over the coming months and produce vaccines which are more effective than the current “emergency brake” Pfizer, Moderna, Oxford, ICMR etc candidates. The maximum vaccination rate possible should also increase significantly over time. So we shall not be surprised if you require two or three trips to the vax clinic over the next couple of years but then cease to require them any longer.

### For policy makers

The set of simulation runs we have shown here is tiny, but hopefully representative. After the initial pace of the epidemic is slowed down, we shall have to shift focus to long-term eradication of the disease, and sponsor research and development drives accordingly. Human trials of vaccines which promise longer-lasting immunity than the current front-runners must continue despite the availability of the current candidates. Parallelly, research on a cure for the disease should also be carried out at full throttle. If an effective antidote can be discovered then that will enable a two-pronged attack on the pathogen. Given the speed with which vaccines were developed, we are hopeful that they may be deployed with the same lightning efficiency to reduce COVID-19 to a nonentity.

## §6 VACCINE HESITANCY

### QUESTION : Myself I am eager for the corona vaccine but I know people who are anti-vaxxers and will refuse it. Should I be concerned about this ?

Vaccine hesitancy, also called vaccine resistance, refers to the unwillingness of some people to receive immunizations. With a disease like COVID-19 where the vaccines have been assembled in a remarkably short time, hesitancy might be higher than usual if people fear that these early vaccines are shoddy jobs. Hesitancy is definitely a challenge to any public health endeavour – the question here is, can it cripple or nullify the COVID-19 elimination efforts, and can the resisters pose a risk to society ?

To address this issue, we expand the basic model to include four classes – high-interaction compliant, high-interaction hesitant, low-interaction compliant and low-interaction hesitant. Let *ρ* be the fraction of people from both interaction classes who are hesitant and let *y*_1_, *y*_2_, *z*_1_ and *z*_2_ denote case counts in the four classes respectively. Some cases among vaccine resisters are only to be expected – a more adverse consequence of hesitancy will be when the resisters transmit the disease to people who are compliant. The dangers are far greater if immunity is **temporary** rather than permanent – in this case a resister harbouring the disease might transmit it to a vaccinee who has been immunized sufficiently long ago. So, when building the model, we take the immunity to be temporary.

The vaccination equations (5,6) remain almost as they were except that the drives now stop when cases plus vaccinees equals 1−*ρ* of the total population – the remaining *ρ* watch the vax drive from the sidelines. Hence the new stopping conditions are that *y*_1_+*v* = (1−*ρ*)*N*_1_ and *z*_1_+*w* = (1−*ρ*)*N*_2_. As for the case equations, following the steps leading to the model derivation in each Section, we get for the compliant high-interaction people

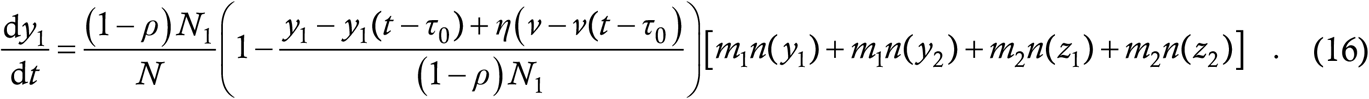

This is almost a carbon copy of (15); the only changes are that the number of compliant high-interaction people is now (1−*ρ*)*N*_1_ instead of *N*_1_ and the counting terms must include all the classes of at large cases. For the hesitant high-interaction class we have

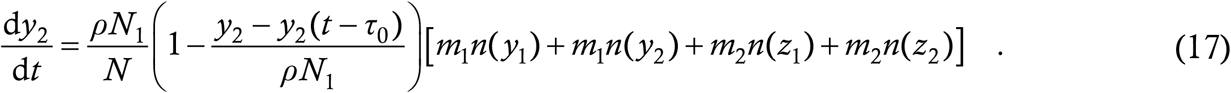

Similarly, for the two remaining classes we have

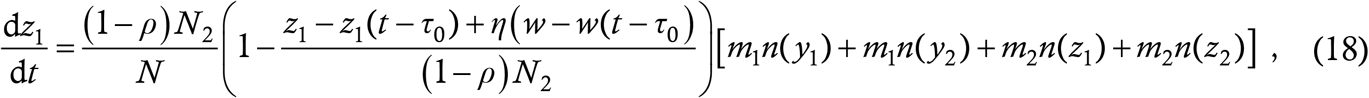

and

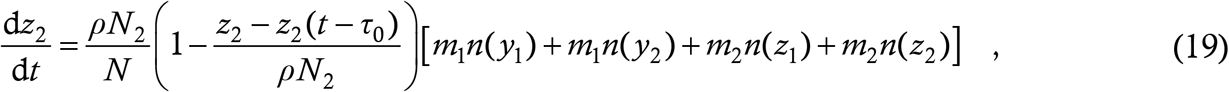

which completes the formulation of the model.

For the simulations, we want situations where the presence of resisters can cause serious trouble, especially cases among the compliants. The natural region of parameter space to look for such situations is near the boundary of the one-shot/ multiwave solutions of the previous Section. Adding a bit of hesitancy to a borderline one-shot solution might destabilize it into a multiwave solution. Accordingly, we again take the 90—10 social structure, 80 percent vaccine effectivity and *τ*_0_ = 200 days, *m*_1_ = 0·70 and *m*_2_ = 0·14 from Figs. 8-10.

With *β* = 0·1, the transition from multi-wave to one-shot solution occurred at *α* = 380 approximately. We go 10 percent above the cutoff to *α* = 420 and see what is the minimum *ρ* required to generate the multiple waves. We find that it is 0·30 i.e. even marginally above the *α* cutoff, a full 30 percent of the population have to be vaccine resisters for the solution to be destabilized. Overcoming 50 percent hesitancy requires an *α* just above 500 – still eminently doable. We are not showing figures here since they don’t differ qualitatively from those of the previous Section.

We now turn to the other simulated situation of the last Section i.e. Figs. 11,12 – 350-day immunity and gradual reopening. With *β* = 0·1, *α* = 720 was the critical value. At *α* = 750 itself, we find we can accommodate 20 percent resisters; at *α* = 1000, more than 50 percent resisters. Another interesting observation is the number of cases accumulated in the compliant group at the end of the outbreak with different proportions of resisters. At *α* = 800, we find 3521 cases within the compliant group with zero resistance, 2985 with 10 percent resistance, 2477 with 20 percent resistance, 1998 with 30 percent resistance and 1544 with 40 percent resistance. Further increasing the proportion of resisters destroys the one-shot elimination solution and the case counts in both groups balloon. The **decrease in compliant cases with increasing hesitancy** might appear a surprise, even a fallacy of our model. It has an easy explanation though – so long as the resisters are not numerous enough to overturn the elimination solution, the total number of cases across all groups increases only marginally with increase in *ρ*. Then, a higher *ρ* means that a greater fraction of the total occurs within this group. In other words, the presence of more resisters ensures that the vaccines go faster to the people who want them, and they are spared from contracting the virus.

In summary, should you be concerned about vaccine hesitancy ?

### ANSWER : We say no

Given the way that corona has brought our world to a standstill, it is very likely that most people will jump at the opportunity of getting a vaccine – 50 percent hesitancy or more appears improbable. And we just showed that small, or not quite small, fractions of resisters do not adversely affect the overall case count. Of course we cannot give a guarantee – it can happen that the authorities are operating so close to the boundary that an elimination effort gets destabilized by the resisters. But that is very unlikely. If your friend is a resister, you can be marginally concerned on his/her behalf since, by resisting, your friend runs a somewhat higher risk of catching corona. However, there is little cause for concern on your own behalf on account of a minority of anti-vaxxers. If anything, their refusal will ensure that the vaccine comes to you faster.

### For policy makers

Vaccine hesitancy is a phenomenon of concern in general but our analysis does seem to indicate that it is one of the least important factors affecting the spread of COVID-19. Of course, our calculation can be challenged in that we sampled only very few data points in an infinite-dimensional parameter space. This challenge is legitimate, but in our defence, we chose these points to be ones where trouble was most likely. If adding resisters to a marginally stable situation doesn’t derail the equilibrium, then it is highly unlikely that adding them to a strongly stable elimination will topple that. Of course, many more simulation runs are needed before implementing policy. But unless there is significant hesitancy, which is very unlikely, this can be one of the lower priority items on the public health authorities’ list. Introducing instant relaxation of restrictions for vaccinees, as discussed in §3, should further reduce people’s tendency to refuse the vaccine. Thus, it seems that vaccine hesitancy can be relegated to the loop line for now. Universities and employers also probably need not worry too much about this phenomenon and should not need to make the vaccine mandatory – if elimination is feasible, it should happen even without this policy.

## §7 CONCLUSION

With the rollout of the first COVID-19 vaccines in the democratic world, questions regarding the fate of the disease are on everyone’s mind. Here we have taken five such questions and attempted to answer them using our in-house mathematical model. Before proceeding further, we wish to repeat that any mathematical model is at best an approximation of reality, and can yield incorrect predictions. While we believe that the DDE model used here is very realistic on account of its transparent philosophy and its use of phenomena-dependent parameters, there is no Law of Nature stating that the evolution of COVID-19 pandemic is indeed governed by (1-19). Thus, our predictions are indicative only – the probability might be greater than even that they come true but that’s the best we can say.

We now describe some of the limitations specific to this model. The two-component interaction-structuring is one of them – it may be that reality is much better modelled by a three- or six-component model. Data obtained from contact tracing drives and mobility studies or surveys should cast insight into this aspect. A minor limitation is that the assumption inherent in (5,6) results in an overcounting of cases as discussed in §1 and §4; as the latter Section shows, the error is small and occurs on the side of caution. A limitation common to all compartmental or lumped parameter models is that they break down when the number of cases involved is small. Thus, “elimination” or “termination” in our model might well correspond to a simmering level in reality with single digit cases being detected each day. In view of the implications of our results on society and public policy, we invite replication of the results using agent-based modeling as well. These models can have very significant predictive power, and statements validated by two independent approaches have even more credibility. Moreover, agent-based model is the only approach which can allow an in-depth exploration of the tail end of the epidemic.

Despite the above limitations however, our model has yielded answers which are qualitatively intuitive. Wherever possible, we have provided logical justification or motivation for our solutions. Preferential vaccination of the high-interaction class is a strategy which is common-sensical and is supported by all the analyses we have performed – in every single situation we have found a significant gain arising from this strategy relative to a proportional vaccination strategy. An equally robust conclusion appears to be the clearance of vaccinated people to normal life – that is probably the easiest way of transitioning from the current state back to a normal state, and one that involves the least risk. Once the vaccination drive starts in earnest, it is probably a question of another four to five months before corona can finally recede from the front and centre of our collective mind. It is too early yet to say whether we shall need repeat trips to the vaccination clinic but for a while at least, that might be on the cards. Research into more effective and long-lasting vaccines as well as antiviral drugs should and hopefully will continue until the menace is completely conquered.

In conclusion, this is probably the last major COVID-19 modeling Article that our group shall write. While we definitely look forward to more work on infectious diseases, we do hope that our next venture can feature a different and more benign disease, that situations don’t crop up with this virus which require yet more pages of analysis and frantic decisions. The conquest of coronavirus, though now in sight, is by no means an indicator of the invincibility of the human race against the forces of Nature. Between the emergence of this virus and its future disappearance, there have been far too many deaths, and far too much economic and social distress for us to feel complacent. The best we can do is use our experience with COVID-19 as a lesson in how we should and should not interact with our surroundings. While a vaccine can deal with one disease, we can take care of all future outbreaks as well as a host of other problems such as catastrophic weather events by living in greater harmony with our environment. Let us hope that the adverse experience of this entire year at least acts as a catalyst in the development of a future, improved way of life.

## Data Availability

The manuscript does not use any data.

## Notes

### Competing Interest Statement

The authors have declared no competing interest.

### Funding Statement

We have not received any funding for this study.

### Author Declarations

No IRB approval required for mathematical modeling study..

